# A Temperature-Dependent Multi-Serotype Model for Evaluating Dengue Vector Control Strategies in Thailand

**DOI:** 10.64898/2026.04.18.26351163

**Authors:** Supassorn Aekthong, Pikkanet Suttirat, Naruemon Rueangkham, Sudarat Chadsuthi, Dominique J. Bicout, Peter Haddawy, Myat Su Yin, Saranath Lawpoolsri, Charin Modchang

## Abstract

**Background:** Dengue remains a major public health challenge in Thailand despite decades of vector control implementation. While mathematical models have explored dengue transmission dynamics, systematic evaluation of current control strategies under realistic operational conditions remains limited.

**Methods:** We developed a temperature-dependent, multi-serotype dengue transmission model that explicitly incorporates three primary vector control strategies: reduction in mosquito biting rates through personal protection measures, further reduction in mosquito birth rates beyond current larval control efforts, and further increase in adult mosquito mortality beyond current adulticide application levels. Using Approximate Bayesian Computation with Sequential Monte Carlo (ABC-SMC), we fitted the model to dengue hemorrhagic fever (DHF) surveillance data from nine province-year combinations representing high (Rayong), moderate (Ratchaburi), and low (Phrae) transmission settings across three years (2006, 2015, and 2017). The model accounts for four dengue serotypes, temperature-dependent mosquito dynamics, and temporary cross-protective immunity between serotypes.

**Results:** The model closely reproduced observed monthly DHF case counts across all nine province-year combinations. Estimated reporting proportions ranged from 1.4% to 16.7%, with the highest values occurring in high-transmission provinces during the 2015 outbreak year. When each strategy was independently intensified by 50% relative to fitted baseline levels, reducing mosquito biting rates and increasing adult mosquito mortality consistently produced greater reductions in transmission than reducing mosquito birth rates. In the highest-transmission scenario (Rayong, 2015), a 50% reduction in biting rate from the baseline level yielded a 96.4% reduction in cumulative infections (95% CrI: 95.4-97.3%), compared with 94.3% (95% CrI: 91.8-95.6%) for a 50% increase in adult mosquito mortality and 77.0% (95% CrI: 58.6-84.6%) for a 50% reduction in mosquito birth rate. Analysis of the time-varying reproduction number (*R*_*t*_) confirmed that interventions targeting adult mosquito-human contact achieved the greatest sustained epidemic suppression, although the relative ranking between bite prevention and adulticide application varied by epidemiological setting.

**Conclusions:** Under the uniform 50% intensification scenario tested, interventions that directly disrupt adult mosquito-human contact, whether through personal protection or adulticide application, substantially outperformed larval control in reducing dengue transmission across diverse Thai settings. These findings support prioritizing personal protection and adulticide application, while the generalizability of this ranking to other intensification levels and settings warrants further investigation.

## Introduction

Dengue fever represents one of the most significant mosquito-borne viral diseases globally, with an estimated 390 million infections occurring annually across more than 100 endemic countries in tropical and subtropical regions [1-3]. The disease is caused by four distinct serotypes of the dengue virus (DENV-1 to DENV-4) and is transmitted to humans through the bites of infected *Aedes* mosquitoes, primarily *Aedes aegypti* and *Aedes albopictus* [2, 4, 5]. Clinical manifestations range from asymptomatic infection or mild undifferentiated fever to dengue fever (DF), and in severe cases, to life-threatening conditions such as dengue hemorrhagic fever (DHF) or dengue shock syndrome (DSS) [2, 5, 6]. Severe dengue can lead to plasma leakage, hemorrhage, and organ impairment, resulting in substantial morbidity and mortality [6, 7].

Thailand remains a persistent hotspot for dengue transmission, with its first reported outbreak of dengue hemorrhagic fever (DHF) in 1958 [8-10]. Dengue incidence fluctuates both geographically and temporally, often exhibiting synchronized timing across provinces [11]. The biological characteristics of the dengue vector are strongly influenced by temperature, with development constrained to a specific optimal range [12-14], while the seasonal peak of dengue tends to coincide with the monsoon period [8]. *Aedes aegypti* and *Aedes albopictus* are the primary vectors responsible for the transmission of dengue in Thailand [15, 16]. Despite decades of vector control implementation, dengue remains a major public health burden in several parts of the country [17, 18], underscoring the continued importance of mosquito population management in reducing transmission risk [17].

Dengue control efforts by the Thai Ministry of Public Health (MoPH), initiated in the 1960s, have primarily relied on vector control measures, including insecticide spraying targeting adult mosquitoes and temephos application for larval control [18]. The 2021 MoPH guidelines recommend four main strategies: physical methods (e.g., environmental management, ovitraps, and personal protection against mosquito bites), biological methods (e.g., larvivorous fish and bacteria), chemical methods (e.g., temephos, insect growth regulators, insecticide-treated nets, and space spraying), and genetic methods [19]. Among these, space spraying—including thermal fogging and ultra-low volume (ULV) spraying—is commonly deployed to reduce adult mosquito populations during outbreaks [19]. However, prolonged insecticide use can drive the development of resistance in mosquito vectors [20], and chemical control should therefore be applied in conjunction with other methods as part of an integrated approach [17, 19].

Effective dengue control in Thailand also depends on timely surveillance. After an outbreak occurred in 1958, a national reporting system for severe forms of dengue, namely dengue hemorrhagic fever (DHF) and dengue shock syndrome (DSS), was established in 1972 and became fully operational by 1974 [21]. The current system integrates epidemiological, virological, and entomological data to detect outbreaks, guide control efforts, and evaluate interventions [22, 23]. However, surveillance often underestimates the true burden due to underreporting of mild cases and incomplete data [24]. Therefore, forecasting systems that predict outbreaks from surveillance data are increasingly essential for proactive dengue control.

Mathematical modeling has increasingly been employed to understand dengue transmission dynamics and evaluate control strategies. Recent models have incorporated temperature-dependent mosquito life history traits to capture seasonal transmission patterns [25], while others have integrated additional climatic factors such as rainfall and humidity [26, 27]. García-Carreras et al. [11] developed a comprehensive multi-serotype model accounting for cross-protection between serotypes and temperature-driven synchronization of epidemics across Thailand. However, a critical gap remains: existing models have not systematically incorporated and evaluated the effectiveness of current vector control interventions under realistic operational conditions.

This study addresses this gap by developing a temperature-dependent, multi-serotype dengue transmission model that explicitly incorporates vector control interventions implemented by the Thai MoPH. We extend the framework of García-Carreras et al. [11] to evaluate three primary control strategies: (1) reduction in mosquito biting rates through personal protection measures, (2) reduction in mosquito birth rates through larval control and source reduction, and (3) increase in adult mosquito mortality through adulticide application. Using Approximate Bayesian Computation with Sequential Monte Carlo (ABC-SMC), we fit our model to DHF surveillance data from nine province-year combinations representing high (Rayong), moderate (Ratchaburi), and low (Phrae) transmission settings across three years (2006, 2015, 2017) with varying outbreak intensities. By estimating reporting proportions and calculating time-varying reproduction numbers (*R*_*t*_), we quantify the differential effectiveness of control strategies across diverse epidemiological contexts. Our findings provide evidence-based insights for optimizing dengue control strategies in endemic settings and highlight the context-dependent nature of intervention effectiveness.

## Methods

### Data

Monthly reported case counts for dengue fever (DF), dengue hemorrhagic fever (DHF), and dengue shock syndrome (DSS) were obtained from the official dengue surveillance system maintained by the Ministry of Public Health (MoPH), Thailand. Although DF and DHF case counts were comparable in the surveillance data, DF cases were considered less reliable due to likely underreporting of milder clinical presentations. We therefore used DHF case counts as the primary outcome for parameter estimation and model fitting.

Monthly minimum and maximum temperatures were obtained from the TerraClimate dataset [28], and the monthly mean temperature for each province was calculated as the arithmetic average of these values. Province-level population data were obtained from the Official Registration Statistics Systems of the Bureau of Registration Administration, Thailand (https://stat.bora.dopa.go.th/).

### Model framework

#### Human population dynamics

The model extends the framework of García-Carreras et al. [11] and explicitly incorporates four dengue virus serotypes (DENV-1 to DENV-4), tracking both primary and secondary infections in the human population. Figure 1 presents a simplified schematic of the compartmental model structure illustrated for DENV-1 only; the full model applies analogous compartments and transitions for all four serotypes with symmetric transmission between humans and mosquitoes.

**Figure 1.**
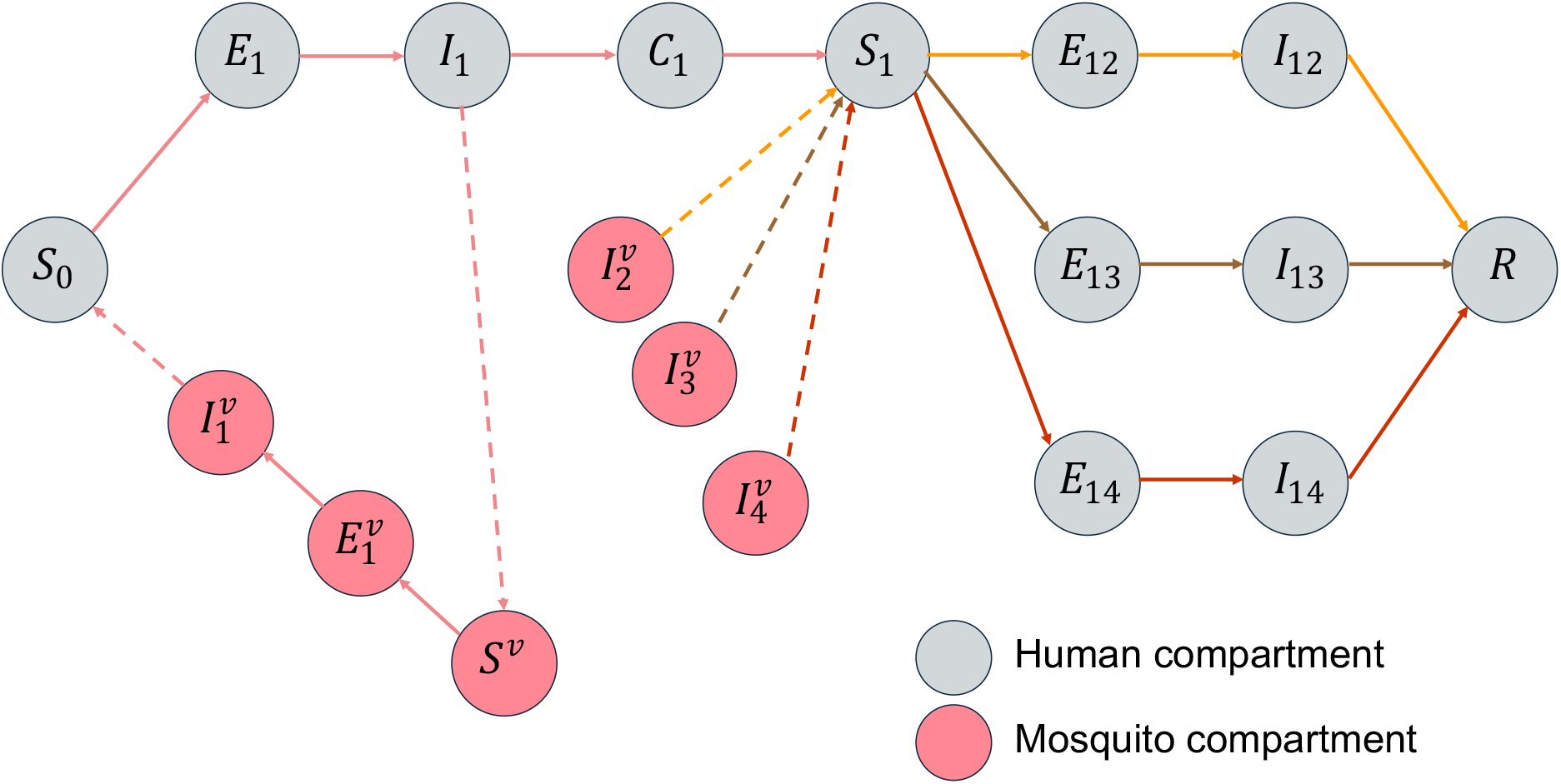
Schematic diagram of the compartmental model framework for dengue transmission. This simplified representation illustrates the model structure for dengue virus serotype 1 only; the full model incorporates four serotypes with symmetric transmission between humans and mosquitoes. In the human population, susceptible individuals (S_0_) become exposed (E_1_) following an infectious mosquito bite. After the intrinsic incubation period, exposed individuals progress to infectious (I_1_) and then develop lifelong immunity to serotype 1 while entering a period of temporary cross-immunity to serotypes 2, 3, and 4 (C_1_). Once cross-immunity wanes, individuals become susceptible to other serotypes (S_1_). Following secondary infection with a different serotype, individuals are assumed to acquire lifelong immunity to all dengue serotypes (R). In the mosquito population, susceptible mosquitoes (S^v^) become exposed after biting an infectious human, then progress to infectious 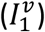 following the extrinsic incubation period. Solid arrows indicate transitions between epidemiological compartments, while dashed arrows represent transmission pathways between human and mosquito populations.

In the human population, individuals begin as fully susceptible to all four serotypes (S_0_). Upon infection with serotype *i*, individuals progress through an exposed state (E_i_) during the intrinsic incubation period and then enter the infectious state (I_i_). Following recovery from primary infection, individuals acquire lifelong immunity to the infecting serotype and enter a period of temporary cross-protective immunity against the remaining three serotypes (C_i_). Once this cross-protection wanes, individuals become susceptible to secondary infection with a heterologous serotype (S_i_). After secondary infection and recovery, individuals are assumed to acquire lifelong immunity to all four serotypes and enter the recovered class (R). The model thus permits a maximum of two sequential dengue infections per individual, consistent with the observation that secondary infections account for the majority of severe dengue cases [29]. Human birth and death rates were set to correspond to an average life expectancy of 77 years, and total population size was held constant throughout each simulation.

Temporal shifts in the predominance of dengue serotypes have been documented across Thai provinces [30, 31], suggesting that serotypes differ in their transmission potential. To capture this heterogeneity, we fixed the transmission rate of DENV-1 as a reference value following [11] and estimated relative transmissibility coefficients for DENV-2, DENV-3, and DENV-4 (denoted *a*_2_, *a*_3_ and *a*_4_) that express each serotype’s transmission rate relative to this baseline. Symmetric transmission between humans and mosquitoes was assumed for each serotype. These relative coefficients are intended to capture differences in transmission potential among serotypes rather than to estimate absolute serotype-specific transmission rates. The dynamics of the human population are governed by the following system of ordinary differential equations

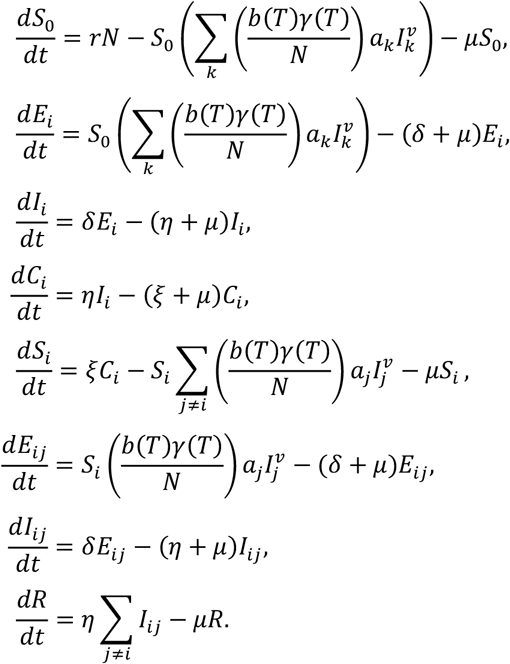

The total human population is partitioned into the following compartments: S_0_ (susceptible to all four serotypes); E_i_ (exposed to serotype *i*); I_i_ (infectious with serotype *i*); C_i_ (recovered from serotype *i* with temporary cross-immunity to other serotypes); S_i_ (susceptible to heterologous serotypes after cross-protection wanes); E_ij_ (exposed to serotype *j* after primary infection with serotype *i*); I_ij_ (infectious with serotype *j* following prior infection with serotype *i*); and R (recovered with lifelong immunity to all serotypes following secondary infection). The total population size is constant and equals the sum of all compartments. Model parameters are summarized in Table 1.

**Table 1.**
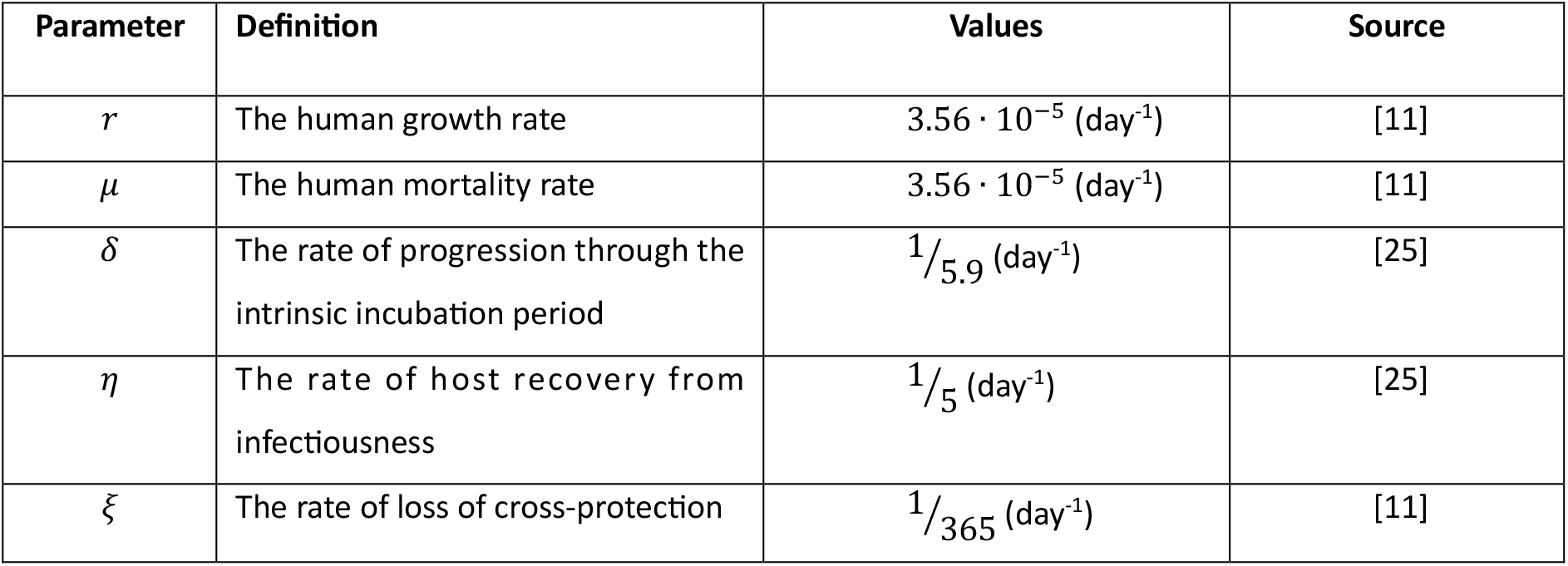
Summary of parameters in the model.

| Parameter | Definition | Values | Source |
| --- | --- | --- | --- |
| $r$ | The human growth rate | $3.56 \cdot 10^{-5} \text{ (day}^{-1}\text{)}$ | [11] |
| $\mu$ | The human mortality rate | $3.56 \cdot 10^{-5} \text{ (day}^{-1}\text{)}$ | [11] |
| $\delta$ | The rate of progression through the intrinsic incubation period | $1/5.9 \text{ (day}^{-1}\text{)}$ | [25] |
| $\eta$ | The rate of host recovery from infectiousness | $1/5 \text{ (day}^{-1}\text{)}$ | [25] |
| $\xi$ | The rate of loss of cross-protection | $1/365 \text{ (day}^{-1}\text{)}$ | [11] |

#### Mosquito population dynamics

The adult mosquito population (*N*^*v*^) is partitioned into susceptible (*S*^*v*^), exposed 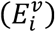, and infectious 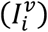 compartments, where the subscript *i* denotes the infecting serotype. Once infected, mosquitoes remain infectious for the duration of their lifespan. Mosquito life history traits—including biting rate, development rate, mortality rate, and the probability of transmission per bite—were modeled as temperature-dependent using fitted thermal response curves reported in previous studies [11, 13, 25]. All mosquito parameter values were based on *Aedes aegypti* [13, 25]. The recruitment rate of susceptible mosquitoes was scaled relative to the human population to ensure consistency across province-level simulations, as the population sizes in this study differ from those in [11]. The dynamics of the mosquito population are governed by the following ordinary differential equations:

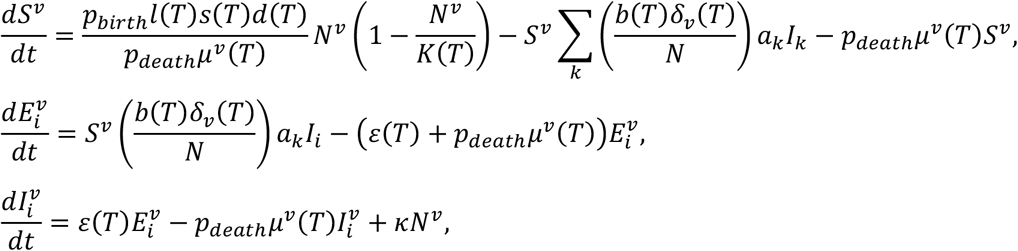

where

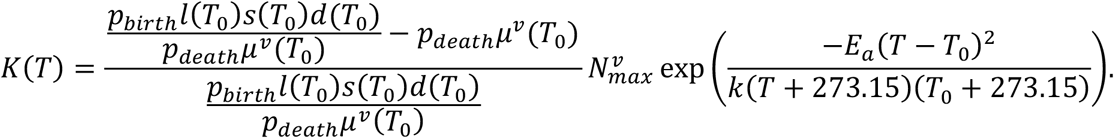

The temperature-dependent mosquito carrying capacity, *K*(*T*) was adapted from [25] and modified to incorporate the effects of the vector control parameters *p*_*birth*_ and *p*_*death*_. Following [11, 25], we set the reference temperature at *T*_0_ = 29 ℃, corresponding to the maximum carrying capacity; the activation energy at *E*_*a*_ = 0.05; the Boltzmann constant at *k* = 8.617 · 10^−5^; and the maximum mosquito-to-human ratio at 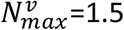.

#### Incorporation of vector control parameters

To represent the vector control interventions implemented by the Department of Disease Control, MoPH, Thailand [15, 19], we incorporated two control parameters into the mosquito population dynamics. The parameter *p*_*birth*_ represents a proportional reduction in the recruitment rate of new susceptible mosquitoes, capturing the combined effects of larval control measures such as larvicide application (e.g., temephos), elimination of breeding sites, container management, and environmental modification. The parameter *p*_*death*_ represents an increase in the mortality rate of adult mosquitoes, reflecting the effects of adulticide interventions including thermal fogging and ultra-low volume (ULV) spraying. Both parameters were estimated as monthly values through ABC-SMC to allow temporal variation in control effort intensity. The description and values of all temperature-dependent mosquito parameters are summarized in Table 2.

**Table 2.** Temperature-dependent mosquito life history parameters. Following [13], each trait of *Aedes aegypti* was fitted to a Brière function, *B*(*T*) = *c*(*T* − *T*_*min*_)(*T* − *T*_*max*_), or a quadratic function, 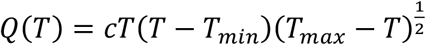, where *T* is the temperature, *c* is the rate constant, *T*_*min*_ is the minimum critical temperature and *T*_*max*_ is the maximum critical temperature. Trait values were set to zero for temperatures outside the critical range [*T*_*min*_ − *T*_*max*_], following [13, 25].

| Trait | Definition | Function | c | $T_{min}$ | $T_{max}$ |
| --- | --- | --- | --- | --- | --- |
| $b$ | The mosquito biting rate (day<sup>-1</sup>) | Brière | $2.02 \cdot 10^{-4}$ | 13.35 | 40.08 |
| $\gamma$ | The probability of mosquito infectiousness | Brière | $8.49 \cdot 10^{-4}$ | 17.05 | 35.83 |
| $l$ | Eggs laid per female per day | Brière | $8.56 \cdot 10^{-3}$ | 14.58 | 34.61 |
| $s$ | The probability of egg-to-adult survival | Quadratic | $-5.99 \cdot 10^{-3}$ | 13.56 | 38.29 |
| $d$ | Mosquito egg-to-adult development rate (day<sup>-1</sup>) | Brière | $7.86 \cdot 10^{-5}$ | 11.36 | 39.17 |
| $\varepsilon$ | The virus extrinsic incubation rate (day<sup>-1</sup>) | Brière | $6.65 \cdot 10^{-5}$ | 10.68 | 45.90 |
| $\mu_v$ | The mortality rate of mosquitoes (day<sup>-1</sup>) | $\frac{1}{\text{Quadratic}}$ | $-1.48 \cdot 10^{-1}$ | 9.16 | 37.73 |
| $\delta_v$ | Probability of mosquito infection | Brière | $4.91 \cdot 10^{-4}$ | 12.22 | 37.46 |

### Vector control strategies

To evaluate the effectiveness of vector control interventions on dengue transmission, we simulated three distinct intervention strategies by modifying specific model parameters while holding all other parameters at their posterior median values from ABC-SMC estimation. The three strategies correspond to the primary operational approaches currently implemented by the Thai MoPH [15, 19]:

#### Strategy 1: Reduction in mosquito-human contact through bite prevention

This strategy represents personal protective measures—including insect repellents, protective clothing, bed nets, and window screens—that reduce effective mosquito–human contact. We modeled this intervention by reducing the temperature-dependent mosquito biting rate, *b*(*T*), by 50% from its fitted baseline value.

#### Strategy 2: Reduction in mosquito recruitment through larval control

This strategy encompasses source reduction measures including elimination of breeding sites, container management, and larvicide application (e.g., temephos) that prevent adult mosquito emergence. We implemented this by reducing the mosquito birth rate parameter *p*_*birth*_by 50% from its posterior median value, which already reflects current larval control efforts.

#### Strategy 3: Increase in adult mosquito mortality through adulticide application

This strategy represents chemical control measures targeting adult mosquitoes, primarily space spraying deployed during outbreaks. We modeled this intervention by increasing the mosquito mortality parameter *p*_*death*_by 50% from its posterior median value, which already incorporates the effects of ongoing adulticide application.

A uniform 50% modification was applied across all three strategies to enable direct comparison of their relative effectiveness at a common intensification level. Importantly, because the baseline parameter values (*p*_*birth*_ and *p*_*death*_) were estimated through ABC-SMC from surveillance data collected during periods of active vector control, these baseline values already reflect existing operational control efforts. The 50% modifications therefore represent additional intensification beyond current practice, rather than interventions applied from a hypothetical no-control state. This moderate effect size was selected to represent a plausible incremental improvement in operational coverage for demonstration purposes, rather than theoretical maximum effects requiring perfect implementation.

### Reporting proportion

Dengue surveillance systems are known to substantially underestimate the true infection burden, with previous studies estimating that reported cases capture only 0.35– 14.7% of actual infections [32-35]. This underreporting reflects multiple factors, including the high proportion of asymptomatic and mild infections, limited healthcare-seeking behavior for non-severe cases, and the predominant capture of hospitalized patients with severe manifestations by surveillance systems.

To account for this surveillance gap, we incorporated a reporting proportion parameter (*ρ*) that links model-simulated infections to observed DHF case counts. Monthly simulated DHF cases were calculated as the sum of new secondary infections across all four serotypes, based on evidence that secondary infections carry elevated risk for severe disease [29]. These simulated secondary infections were then multiplied by ρ, which was estimated simultaneously with other model parameters through ABC-SMC.

The reporting proportion thus represents the composite probability that a secondary dengue infection will: (1) progress to DHF rather than mild or asymptomatic disease, (2) lead to healthcare seeking, (3) be correctly diagnosed, and (4) be captured by the surveillance system. This parameter was estimated independently for each province-year combination, allowing it to capture differences in healthcare accessibility, surveillance intensity, and potentially serotype-specific disease severity patterns. By explicitly estimating *ρ*, our framework enables inference of the total infection burden that underlies the observed case counts, providing a more accurate basis for assessing the true impact of control interventions on dengue transmission.

### Province and year selection

To investigate the effectiveness of vector control strategies across diverse epidemiological contexts, we selected three provinces representing distinct transmission intensities based on their average monthly reported DHF incidence during 2003–2022 (Figure 2a): Rayong (high transmission; mean 9.72 cases per 100,000 population), Ratchaburi (moderate transmission; mean 6 cases per 100,000), and Phrae (low transmission; mean 2 cases per 100,000). For each province, we selected three years—2006, 2015, and 2017—that exhibited varying outbreak intensities within each setting (Figure 2b), yielding nine province-year combinations for analysis.

**Figure 2.**
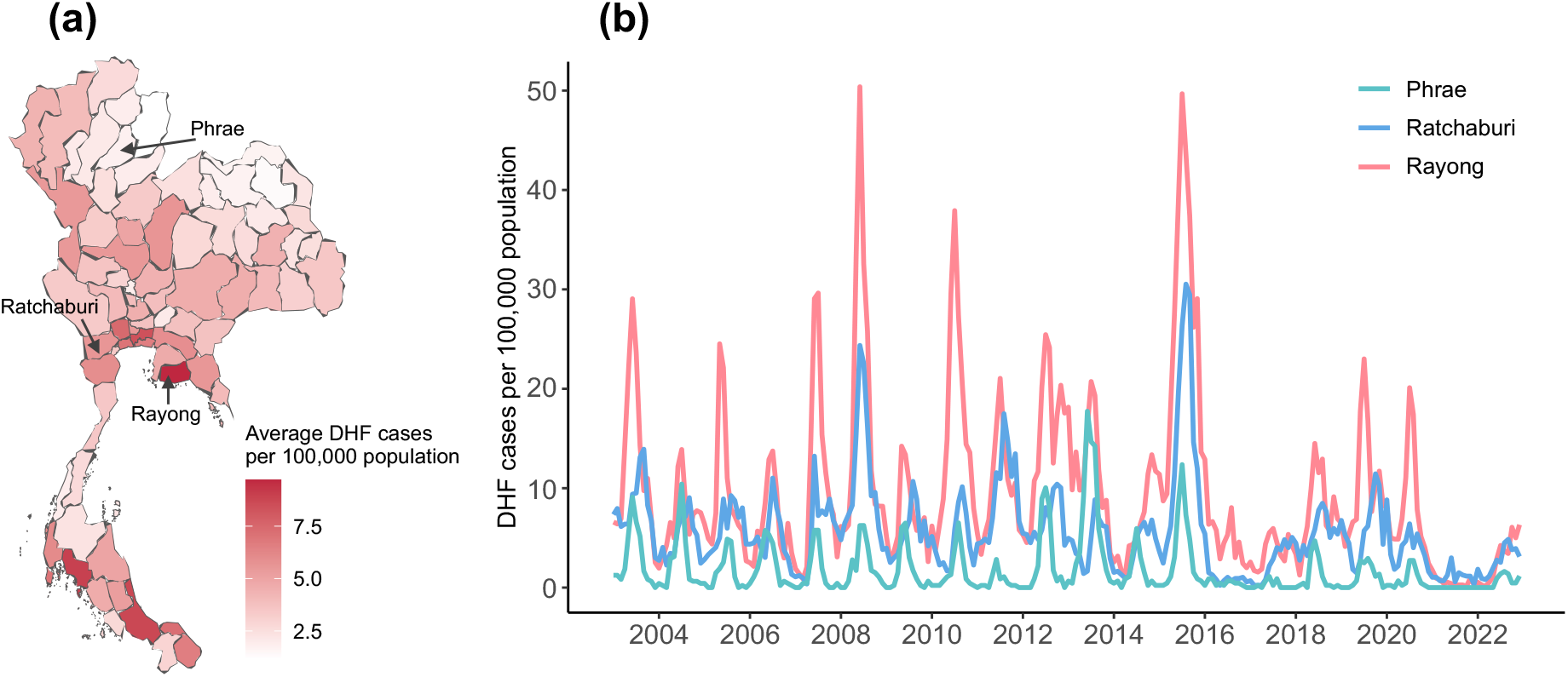
Spatial and temporal patterns of dengue hemorrhagic fever (DHF) incidence in Thailand used for model selection and analysis. **(a)** Spatial distribution of average monthly reported DHF cases per 100,000 population across all provinces in Thailand from 2003 to 2022. **(b)** Time series of average monthly reported DHF cases per 100,000 population in Rayong, Ratchaburi and Phrae.

### Model fitting using Approximate Bayesian Computation with Sequential Monte Carlo

We estimated model parameters using Approximate Bayesian Computation with Sequential Monte Carlo sampling (ABC-SMC) [36], a likelihood-free Bayesian inference method well suited to complex dynamical models for which the likelihood function is analytically intractable or computationally prohibitive. Rather than evaluating the likelihood directly, ABC-SMC compares summary statistics of simulated data with those of the observed data and retains parameter combinations (particles) that produce simulations sufficiently close to the observations.

The ABC-SMC algorithm proceeds through a series of generations, each imposing a progressively stricter acceptance criterion to guide the particle population toward regions of high posterior probability. In the first generation, candidate parameter vectors (particles) are drawn from the prior distributions and used to simulate the model. Particles whose simulated output falls within a specified tolerance of the observed data are retained, while the remainder are discarded. In subsequent generations, a particle is selected from the retained population of the previous generation with probability proportional to its importance weight, then perturbed by a proposal kernel to generate a new candidate. This candidate is accepted only if its simulation meets the tightened tolerance threshold for the current generation. By iteratively reducing the tolerance across generations, the algorithm produces an increasingly refined approximation to the posterior distribution without requiring explicit likelihood evaluation [36].

The model was fitted independently to each of the nine province-year combinations, with 28 parameters estimated simultaneously per fit. Of these, the vector control parameters *p*_*birth*_ and *p*_*death*_ were estimated as monthly values (12 values each per year), the relative serotype transmissibility coefficients (*a*_2_, *a*_3_, *a*_4_) were estimated as annual values, and the reporting proportion (*ρ*) was estimated as a single value per province-year. A total of *N*_*par*_ = 200 particles were retained per generation to balance computational feasibility with adequate posterior characterization given the high dimensionality of the parameter space.

The distance between simulated and observed data was quantified using the root mean square error (RMSE):

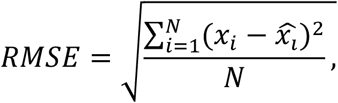

where *x*_*i*_ is the number of reported DHF cases in month 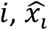 is number of simulated cases in month *i, N* is the number of months in the simulated period. The RMSE thus provides an interpretable measure of the average monthly deviation between simulated and observed case counts.

The initial tolerance threshold for the first ABC-SMC generation was set to half the maximum reported monthly case count in each setting to ensure a sufficient acceptance rate, and the tolerance was subsequently reduced by 15% at each successive generation. The algorithm terminated when the RMSE fell below 14.5% of the mean monthly case count for each setting, or when it reached an absolute threshold of approximately 1 for settings in which the mean monthly case count was fewer than 2. Initial parameter values were drawn from the uniform prior distributions specified in **Table 3**. At each generation, a new proposal was generated by sampling a particle from the previous population with probability proportional to its weight and perturbing it using a truncated multivariate normal kernel centered on the selected particle, with covariance matrix computed from the full previous-generation population.

**Table 3.** Prior distributions for all estimated parameters. The parameter *a*_*i*_ (*i* = 2, 3, 4) denotes the relative transmission coefficient for serotype *i*, with serotype 1 fixed as the reference (*a*_1_ = 1).

| Parameter | Prior distribution |
| --- | --- |
| $a_2, a_3, a_4$ | (0.1,10) |
| $p_{birth}$ | (0.2,1) |
| $p_{death}$ | (1,10) |
| $\rho$ | (0.00001,0.25) |

### Simulation settings and initial conditions

All model simulations were performed in R version 4.4.2 using the “lsoda” solver from the “deSolve” package, which provides an adaptive-step integration method suitable for stiff and non-stiff ODE systems. The duration of temporary cross-protective immunity between serotypes was fixed at 365 days. In the absence of daily temperature data, daily temperatures were approximated by the monthly mean temperature for the corresponding province and year. The total population for each province was set to its average value in 2018.

Initial conditions for the fitting period were obtained through a two-stage procedure. First, the model was pre-equilibrated by running a burn-in simulation until the system reached a dynamic steady state under baseline parameter values. The resulting steady-state proportions of susceptible, exposed, infectious, and recovered individuals were broadly consistent with empirical seroprevalence data from a cohort study in Ratchaburi province [37], providing independent support for the equilibrium conditions. Second, the state variables at the end of the burn-in period were used as fixed initial conditions for the ABC-SMC fitting procedure, thereby eliminating the need to estimate initial conditions as additional free parameters and reducing the dimensionality of the estimation problem.

During model fitting, the monthly simulated DHF cases were computed as the total number of new secondary infections summed over each calendar month, multiplied by the estimated reporting proportion *ρ*. These values were then compared to the observed monthly DHF case counts to compute the RMSE for each particle.

### Estimation of the time-varying effective reproduction number

The time-varying effective reproduction number, *R*_*t*_, quantifies the average number of secondary human infections generated by a single infectious individual at time *t* in a partially immune population [38]. When *R*_*t*_ > 1, the epidemic is growing; when *R*_*t*_ < 1, transmission is declining and the outbreak will eventually subside in the absence of external introductions. *R*_*t*_ therefore serves as a real-time indicator of intervention effectiveness.

We estimated *R*_*t*_ using the “EpiEstim” R package developed by Cori et al. [38], which employs a Bayesian framework to infer *R*_*t*_ from time series of incident cases and a specified generation time distribution. The generation time—defined as the interval between the time of infection of a primary case and the time of infection of a secondary case infected by that primary case—was modeled as a gamma distribution with a mean of 18.2 days and a standard deviation of 6.1 days, consistent with estimates for dengue that account for both the intrinsic and extrinsic incubation periods [31].

For each of the four scenarios (baseline and three vector control strategies), we used the median daily number of new infections (both primary and secondary) from the posterior ensemble as input to EpiEstim. *R*_*t*_ was estimated using a weekly (7-day) sliding window, with estimation commencing from the earliest date at which two conditions were satisfied: (1) the cumulative number of simulated infections exceeded 26 cases, ensuring a target posterior coefficient of variation of 0.2 for the *R*_*t*_ estimate, and (2) at least one mean generation time (19 days) had elapsed since the start of the simulation. To integrate both the magnitude and duration of epidemic control, we additionally calculated the net area difference (Δ*A*) between periods with *R*_*t*_ > 1 and *R*_*t*_ < 1, where negative values indicate successful epidemic suppression. A detailed description of the EpiEstim framework is provided in [38].

## Results

### Model fitting

The fitted models closely reproduced the observed monthly DHF case counts across all nine province-year combinations, successfully capturing both the seasonal timing and magnitude of outbreak peaks. Figure 3 illustrates the model fit for Rayong during 2015, the province-year combination with the highest reported incidence (2,146 DHF cases). The ensemble of 200 parameter sets (particles) retained in the final ABC-SMC generation (grey lines) closely tracked the observed monthly case counts (pink line), with variation among particles reflecting parameter uncertainty. Comparable fitting quality was achieved for all remaining province-year combinations (Figures S1, Supplementary Information).

**Figure 3.**
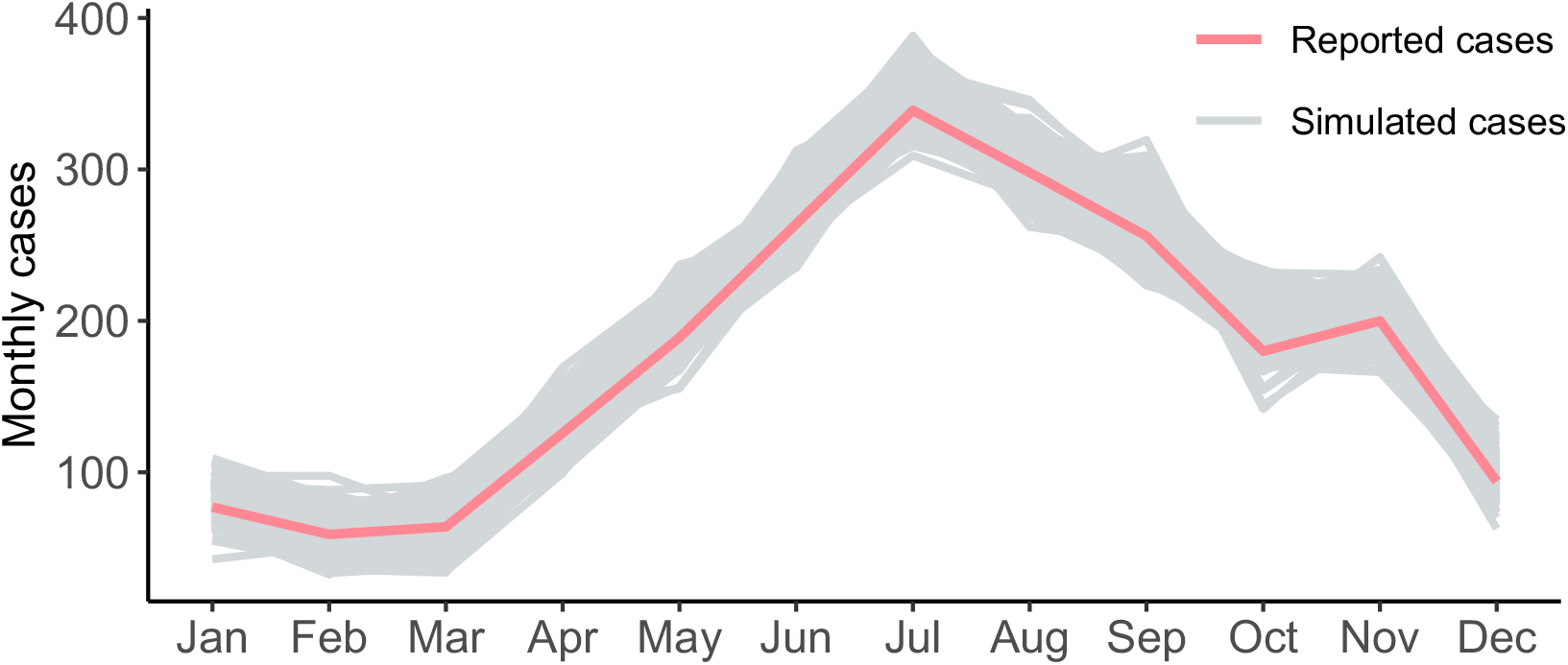
Model fitting to monthly DHF surveillance data in Rayong province, 2015. The pink line shows observed monthly DHF cases (total: 2,146 cases). Grey lines represent the ensemble of simulated monthly DHF cases from 200 parameter sets (particles) retained in the final generation of the ABC-SMC algorithm, with each line corresponding to one parameter combination that satisfied the acceptance criterion (RMSE < 14.5% of mean monthly cases). The simulated DHF cases were calculated as the monthly sum of secondary dengue infections multiplied by the estimated reporting proportion.

### Estimated reporting proportions

The reporting proportion parameter (*ρ*), estimated simultaneously with other model parameters through ABC-SMC, exhibited substantial variation across province-year combinations (Figure 4; Figure S2, Supplementary Information). In Rayong, the estimated *ρ* was 0.037 (95% CrI: 0.024–0.066) in 2006, 0.166 (95% CrI: 0.113–0.212) in 2015, and 0.038 (95% CrI: 0.027–0.057) in 2017. The approximately four-to five-fold higher reporting proportion in 2015 coincided with an exceptionally large outbreak in Rayong (2,146 DHF cases, compared with 324 in 2006 and 432 in 2017), potentially reflecting enhanced case detection or increased progression to severe disease during periods of intense transmission.

**Figure 4.**
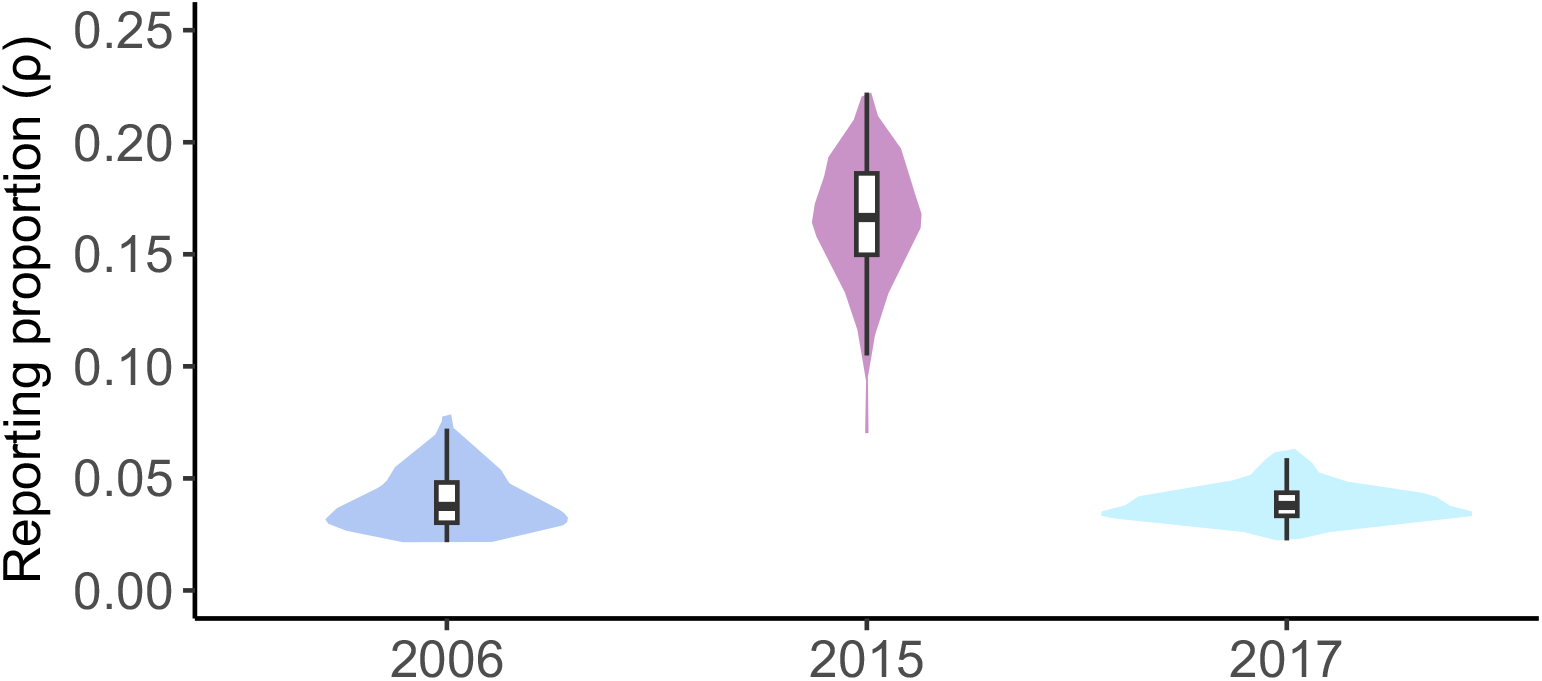
Posterior distributions of estimated reporting proportions for Rayong province across three years (2006, 2015, 2017). Violin plots display the distribution of reporting proportion estimates from 200 parameter sets retained in the final generation of ABC-SMC for each year. The reporting proportion represents the fraction of secondary dengue infections that progress to DHF and are captured by the surveillance system. The width of each violin indicates the kernel density estimate of the posterior distribution, with embedded box plots showing the median (horizontal line), interquartile range (box), and 95% credible intervals (whiskers). The four-fold higher reporting proportion in 2015 (median: 0.166) compared to 2006 (median: 0.037) and 2017 (median: 0.038) coincides with the exceptional outbreak that year (2,146 DHF cases), suggesting enhanced case detection or increased progression to severe disease during periods of intense transmission.

A similar temporal pattern was observed in Ratchaburi, where ρ was elevated during 2015 (0.167, 95% CrI: 0.083–0.245) relative to other years, coinciding with its peak of 1,432 reported DHF cases. In contrast, Phrae—representing a low-transmission setting with only 14– 78 annual DHF cases across the study years—maintained consistently low reporting proportions across all three years (Figure S2, Supplementary Information). Across all nine province-year combinations, estimated *ρ* values ranged from approximately 1.4% to 16.7%, with the highest values concentrated in the high-transmission provinces during the 2015 outbreak year.

### Effects of vector control strategies on transmission dynamics

Figure 5a illustrates the temporal dynamics of daily new infections under the baseline and three intervention scenarios for Rayong during 2015. Under the 50% intensification scenario, all three vector control strategies substantially reduced transmission relative to the baseline, with peak daily infections decreasing from approximately 200 (baseline) to below 100 under each intervention. Under the 50% intensification scenario, Strategies 1 (bite prevention) and 3 (adulticide) demonstrated comparable and pronounced efficacy. Strategy 2 (larval control) yielded the smallest reduction. This hierarchical pattern—Strategies 1 ≈ 3 > Strategy 2—was consistent across all nine province-year combinations at the tested intensification level.

**Figure 5.**
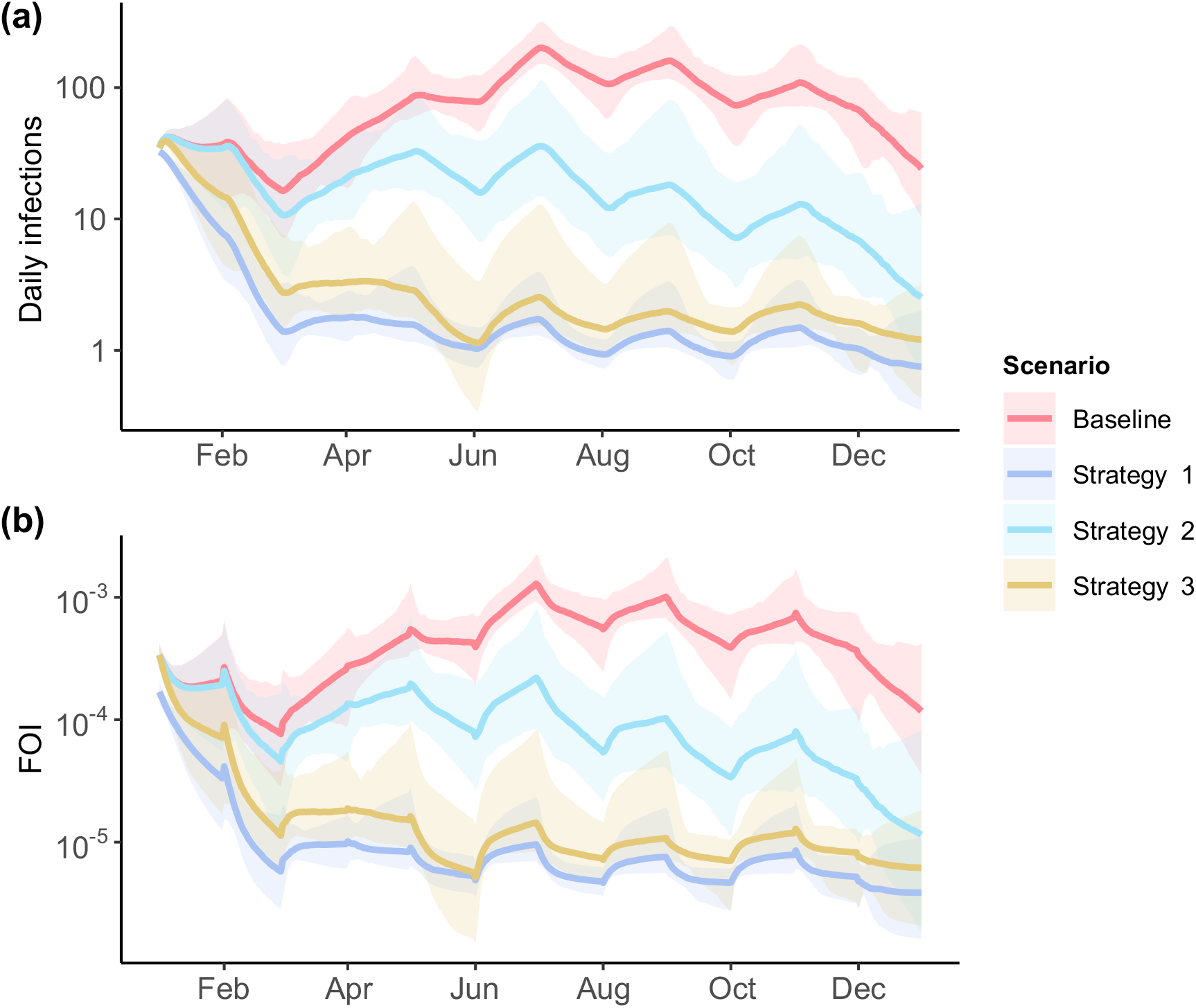
Temporal dynamics of dengue transmission in Rayong province during 2015 under baseline and vector control scenarios. **(a)** Daily number of new dengue infections (primary and secondary) simulated using posterior median parameters from ABC-SMC estimation. Peak baseline transmission reaches approximately 200 daily infections during July. **(b)** Daily force of infection (FOI), representing the per-capita rate at which susceptible individuals acquire infection, calculated as the sum of serotype-specific transmission rates weighted by infectious mosquito abundance. Strategy 1 (purple): 50% reduction in mosquito biting rate, modeling personal protection measures and bite avoidance. Strategy 2 (sky blue): 50% reduction in mosquito birth rate parameter (*p*_*birth*_), representing larvicide application and breeding site reduction. Strategy 3 (sandstone): 50% increase in mosquito mortality rate parameter (*p*_*death*_), simulating adulticide interventions.

The cumulative force of infection (FOI), representing the per-capita rate at which susceptible individuals acquire infection summed across all four serotypes, confirmed the differential impact of the three strategies (Figure 5b). Across all province-year combinations, the FOI followed a consistent hierarchy: baseline > Strategy 2 > Strategy 3 > Strategy 1. Strategy 1 produced the lowest FOI because the biting rate enters the transmission equations quadratically, simultaneously reducing both mosquito-to-human and human-to-mosquito transmission. Strategy 3 reduced FOI by increasing adult mosquito mortality, thereby shortening the mean duration of infectiousness and reducing the probability of surviving through the extrinsic incubation period. Strategy 2 produced the smallest FOI reduction, as it targets mosquito recruitment without directly affecting the existing population of infectious adult mosquitoes.

The impact of each strategy on cumulative infections varied across epidemiological settings (Figure 6b; Figures S3–S10, Supplementary Information). Strategy 1 achieved the largest percentage reduction in cumulative infections in eight of nine province-year combinations, with reductions exceeding 92% in all settings except Phrae 2017 (86.6% reduction). In Rayong 2015, Strategy 1 achieved a 96.4% reduction (95% CrI: 95.4–97.3%), followed by Strategy 3 with 94.3% (95% CrI: 91.8–95.6%) and Strategy 2 with 77.0% (95% CrI: 58.6–84.6%). Strategy 2 exhibited the most variable performance across settings, notably achieving less than 50% reduction in the low-transmission setting of Phrae during 2006 and 2017, where baseline transmission was already minimal (14–78 reported DHF cases annually).

**Figure 6.**
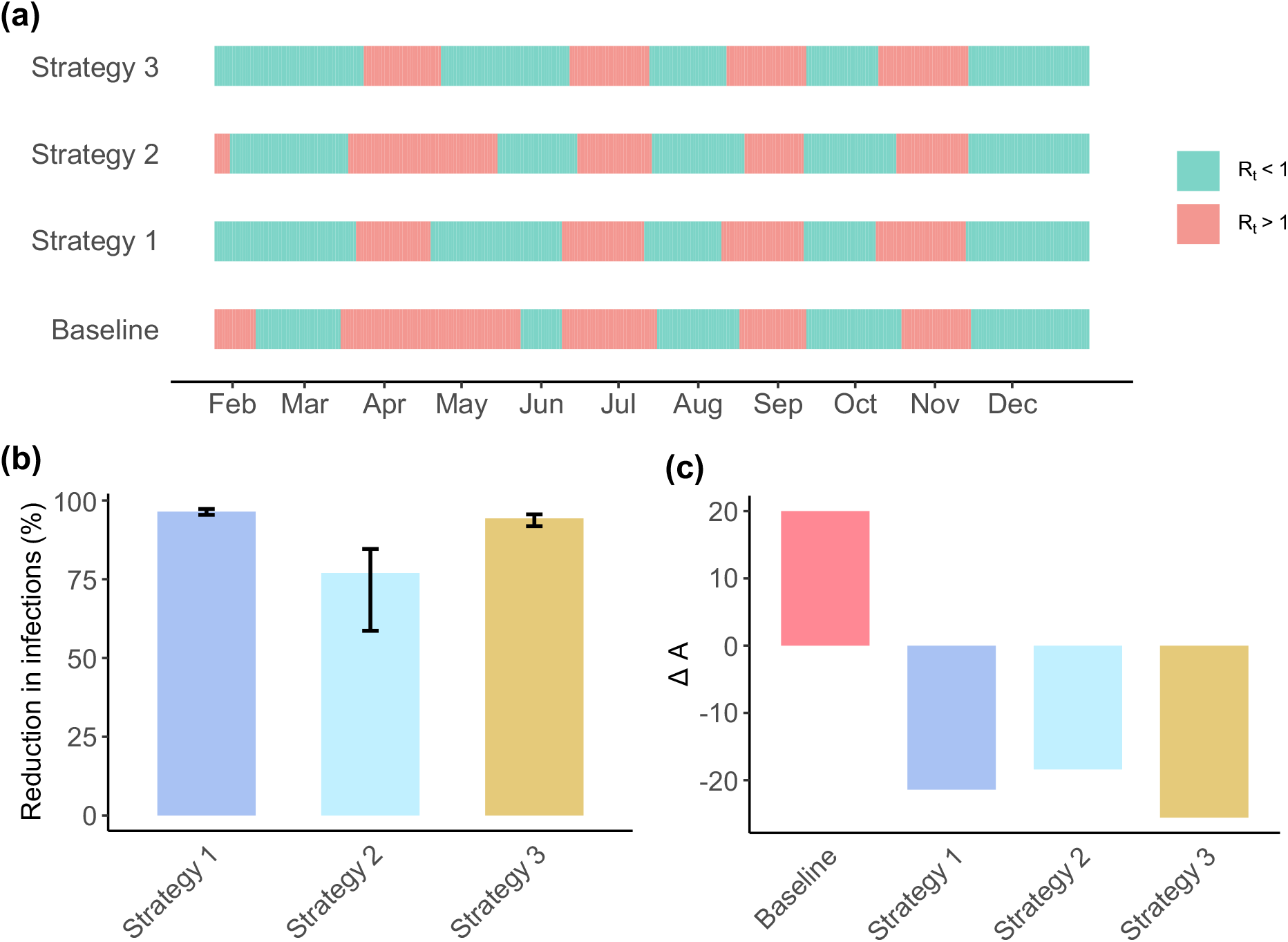
Quantitative assessment of vector control effectiveness on dengue epidemic dynamics in Rayong province, 2015. (a) Time-varying effective reproduction number (*R*_*t*_) estimated using EpiEstim with a weekly sliding window and gamma-distributed generation time (mean: 18.2 days, SD: 6.1 days). Red shading indicates periods of epidemic growth (*R*_*t*_ > 1) and green shading indicates epidemic decline (*R*_*t*_ < 1). The baseline scenario exhibits *R*_*t*_ > 1 for 51.6% of the simulation period, reduced to 37.5% (Strategy 1), 42.2% (Strategy 2), and 37.2% (Strategy 3) under interventions. (b) Percentage reduction in cumulative dengue infections relative to baseline, calculated from 200 parameter sets retained from ABC-SMC. Error bars represent 95% credible intervals derived from the posterior distribution. Strategy 1 (bite prevention): 96.4% reduction; Strategy 3 (adulticides): 94.3% reduction; Strategy 2 (larvicides): 77.0% reduction. (c) Net epidemic intensity (Δ*A*) calculated as the difference between areas above and below the *R*_*t*_ = 1 threshold. Negative values indicate successful epidemic suppression, with larger absolute values representing stronger control. Strategy 3 demonstrates the most effective sustained control (most negative Δ*A*).

### Effectiveness of control measures as reflected in the time-varying reproduction number

Figure 6a presents the temporal evolution of *R*_*t*_ in Rayong during 2015, with periods of epidemic growth (R_*t*_ > 1, red shading) and decline (R_*t*_ < 1, green shading) clearly delineated. Under baseline conditions, R_*t*_ exceeded the epidemic threshold for 51.6% of the simulation period, with sustained epidemic growth during April–May. All three vector control strategies compressed these epidemic periods: Strategy 3 reduced the proportion of time with R_*t*_ > 1 to 37.2%, Strategy 1 to 37.5%, and Strategy 2 to 42.2%. Both Strategies 1 and 3 notably suppressed the April–May transmission peak that dominated the baseline scenario.

The net area difference (ΔA), which integrates both the magnitude and duration of epidemic control relative to the R_*t*_ = 1 threshold, provided a complementary summary measure of overall epidemic suppression (Figure 6c). Strategy 3 yielded the most negative ΔA in the majority of province-year combinations, marginally outperforming Strategy 1 in sustained epidemic suppression, despite Strategy 1 generally achieving larger percentage reductions in cumulative infections. The baseline scenario yielded positive ΔA values across all settings, confirming sustained net epidemic growth in the absence of additional interventions. Taken together, these results indicate that, at the 50% intensification level tested, Strategies 1 and 3 consistently outperformed Strategy 2 across settings, whereas the ranking between Strategies 1 and 3 depended on the specific epidemiological context.

## Discussion

In this study, we developed a temperature-dependent, multi-serotype dengue transmission model that extends the framework of García-Carreras et al. [11] by explicitly incorporating the vector control interventions currently implemented by the Thai Ministry of Public Health. By analyzing nine province-year combinations spanning high (Rayong), moderate (Ratchaburi), and low (Phrae) transmission settings across three years (2006, 2015, 2017), our model provides a comprehensive evaluation of control strategy effectiveness under diverse epidemiological contexts.

Our model assumes that individuals can experience a maximum of two dengue infections, with secondary infections contributing to the simulated DHF cases. This assumption aligns with established evidence that secondary dengue infections carry elevated risk for severe disease manifestations, including DHF and DSS [29]. To reconcile model outputs with surveillance data, we estimated province- and year-specific reporting proportions that align monthly simulated DHF cases with observed reports. Our estimated reporting proportions (ranging from 1.4% to 16.7%) are broadly comparable to those reported in previous studies [32-35], which documented reporting rates between 0.35% and 14.7%, though our upper estimates during the 2015 outbreak year exceeded this range.

The highest estimated reporting proportions occurred in Rayong and Ratchaburi during 2015, coinciding with an exceptional dengue outbreak that resulted in 2,146 and 1,432 reported DHF cases, respectively. This pattern reflects the hyperendemic nature of dengue transmission in these provinces. Rayong’s persistent co-circulation of multiple serotypes [39] and Ratchaburi’s high seroprevalence, with 98% of adults over 25 years showing evidence of prior dengue exposure in a 2012-2015 cohort study [37], underscore the intense transmission in these settings. Notably, while reporting proportions peaked in 2015 for both provinces, they remained substantially lower in 2006 and 2017, suggesting year-to-year variability in surveillance capture or disease severity patterns. In contrast, Phrae exhibited consistently low reporting proportions across all years, corresponding with its classification as a low-transmission setting with only 234 total DHF cases reported across the three study years.

Additionally, serological surveys in Rayong demonstrate age-dependent increases in multitypic exposure, with approximately 60% of 18-year-olds had antibodies to multiple serotypes [40]. This high background immunity suggests that substantial secondary infections occur but remain undetected by surveillance systems. Supporting this interpretation, a Taiwan cohort study found that only 7.8% of secondary dengue infections progressed to severe disease, with risk particularly elevated when inter-infection intervals exceeded two years (OR 3.19, 95% CI: 2.04-5.00) [41].

These findings highlight that routine surveillance captures only a fraction of dengue transmission events, predominantly those resulting in severe disease requiring hospitalization. Our modeling framework could therefore provide a valuable tool for estimating the hidden burden of dengue infections that contribute to ongoing transmission but escape detection through standard surveillance mechanisms. This capability is particularly important for evaluating the true impact of control interventions and planning resource allocation in dengue-endemic settings.

### Impact of vector control strategies on dengue transmission

Our model incorporates all four dengue virus serotypes to estimate the cumulative daily force of infection (FOI) under different vector control scenarios. This comprehensive approach enables quantification of how specific interventions modify overall transmission intensity across serotypes. Across all nine province-year combinations analyzed, vector control interventions consistently reduced the daily FOI relative to baseline conditions.

Among the three control strategies evaluated at the 50% intensification level, reducing the mosquito biting rate achieved the greatest FOI reduction, followed by increasing adult mosquito mortality rates, while reducing mosquito birth rates showed the smallest impact. These modeled interventions correspond directly to established control measures implemented by the Thai Ministry of Public Health [19]. Biting rate reduction represents personal protective measures (e.g., repellents, bed nets, protective clothing), increased mortality reflects adulticide applications (e.g., spraying, and fogging), and birth rate reduction corresponds to larvicidal treatments and source reduction strategies.

The superior efficacy of biting rate reduction might be due to its direct disruption of human-mosquito contact, thereby reducing both mosquito-to-human and human-to-mosquito transmission pathways. This dual effect manifests as substantial reductions in total dengue infections across simulations. Specifically, a 50% reduction in mosquito biting rates from baseline levels achieved >92% reduction in total infections for eight of nine scenarios. The single exception, Phrae 2017, showed an 86.6% reduction, which likely reflects the extremely low baseline transmission in this setting (14 reported DHF cases).

### Effectiveness of control measures as reflected in the time-varying reproduction number

To quantify the epidemiological impact of vector control interventions, we analyzed their effects on the time-varying reproduction number (*R*_*t*_) using two complementary metrics: the proportion of time *R*_*t*_ remained above the epidemic threshold (*R*_*t*_ > 1) and the area difference under the *R*_*t*_ curve (ΔA) relative to baseline. These metrics capture both the duration and intensity of epidemic transmission under different control scenarios.

At the 50% intensification level tested, the relative effectiveness of control strategies varied across province-year combinations, revealing context-dependent intervention impacts. In Rayong 2015, increasing adult mosquito mortality rates (Strategy 3) achieved the most substantial transmission suppression, producing both the shortest epidemic period (lowest proportion of time with *R*_*t*_ > 1) and the greatest reduction in epidemic intensity (most negative ΔA). Reducing mosquito biting rates (Strategy 1) also demonstrated notable efficacy, substantially shortening the epidemic period and moderately reducing intensity. However, this pattern reversed in Rayong 2006 and 2017, where biting rate reduction outperformed mortality rate increases across both metrics. For Ratchaburi and Phrae across all years, increasing mortality rates consistently achieved superior transmission control, followed by biting rate reduction. Notably, reducing mosquito birth rates (Strategy 2) showed limited effectiveness across all scenarios.

The consistently poor performance of birth rate reduction strategies (larvicides and source management) in our simulations aligns with empirical evidence of limited field effectiveness. A systematic review and meta-analysis found that larval control interventions, including container covers, waste management, and breeding site elimination, produced only marginal impacts on *Aedes* populations, with no significant differences among approaches [42]. Implementation gaps further compromised effectiveness, as demonstrated in Myanmar where increased larval control efforts between 2012-2015 failed to meaningfully reduce dengue risk due to insufficient coverage of households, schools, and breeding sites [43]. With the exception of successful programs in Cuba and Singapore, community-based source reduction has achieved limited success, primarily due to inadequate participation and sustainability [44].

Our results demonstrate that interventions targeting adult mosquito-human contact, whether through biting rate reduction or increased adult mortality, substantially outperform larval control in disrupting dengue transmission dynamics. This finding has important implications for resource allocation and intervention prioritization in dengue-endemic settings, suggesting that, at the intensification level modeled here, control programs would benefit from emphasizing adult vector control and personal protection measures, while recognizing that the optimal strategy mix may differ at other intensification levels and that operational and biological constraints may limit field effectiveness. The context-dependent variation in optimal strategies across provinces and years further underscores the need for adaptive, locally tailored intervention approaches rather than uniform national strategies.

### Study Limitations

Several assumptions in our model framework may influence the generalizability of our findings. First, we limited individuals to a maximum of two dengue infections with lifelong immunity following secondary infection, though tertiary and quaternary infections have been documented in hyperendemic settings [45, 46]. This simplification may underestimate transmission dynamics in populations with high force of infection where multiple reinfections contribute to onward transmission. Second, although it has been found that temperature dominates dengue transmission in Thailand [47-51], excluding other potentially important climatic factors—such as rainfall patterns and extreme weather events—that influence mosquito breeding site availability may affect model predictions in settings where these factors play a substantial role. Additionally, we assumed symmetric transmission rates between humans and mosquitoes for each serotype and expressed serotypes 2-4 transmission rates relative to serotype 1, preventing estimation of absolute serotype-specific transmission parameters. The model also lacks spatial structure and human mobility patterns, which are known to influence dengue transmission dynamics [52-56].

Furthermore, our evaluation of vector control strategies employs simplified representations that assume uniform coverage, and consistent effectiveness. The model does not account for the evolution or spatial heterogeneity of insecticide resistance, which could substantially alter intervention effectiveness over time. Moreover, our evaluation was conducted at a single intensification level (50% modification from fitted baseline values), and the relative ranking of control strategies may differ under alternative intensification scenarios. Finally, limiting our analysis to nine province-year scenarios from three provinces may not fully capture the diversity of dengue transmission settings across Thailand’s 77 provinces, each with unique ecological, demographic, and epidemiological characteristics. These limitations suggest that while our model provides valuable insights into the relative effectiveness of control strategies at the intensification level tested, translating these findings into operational guidance requires site-specific validation, and integration with local entomological and resistance monitoring data.

## Data Availability

All data produced in the present study are available upon reasonable request to the authors.

## Acknowledgements

This research has received funding support from The Rockefeller Foundation under the Strengthening the Early Warning and Outbreak Detection Systems through Nation-wide Event-based and Syndromic Surveillance (STONES) project (Grant No. 2021PPI005).

## Competing interests

The authors declare no competing interests.

## Ethics statement

This study received an exemption from ethical review by the Ethics Committee of the Faculty of Tropical Medicine, Mahidol University, Thailand (Documentary Proof of Exemption: MUTM-EXMPT 2024-005, dated 8 July 2024). The exemption was granted as this research involved retrospective analysis of anonymized, aggregated surveillance data with no direct human subject involvement. The Ethics Committee certified that the study complies with the Declaration of Helsinki, ICH Guidelines for Good Clinical Practice, and other international guidelines for human research protection.

## Notes

### Competing Interest Statement

The authors have declared no competing interest.

