## Supplementary Information for "A Temperature-Dependent Multi-Serotype Model for Evaluating Dengue Vector Control Strategies in Thailand"

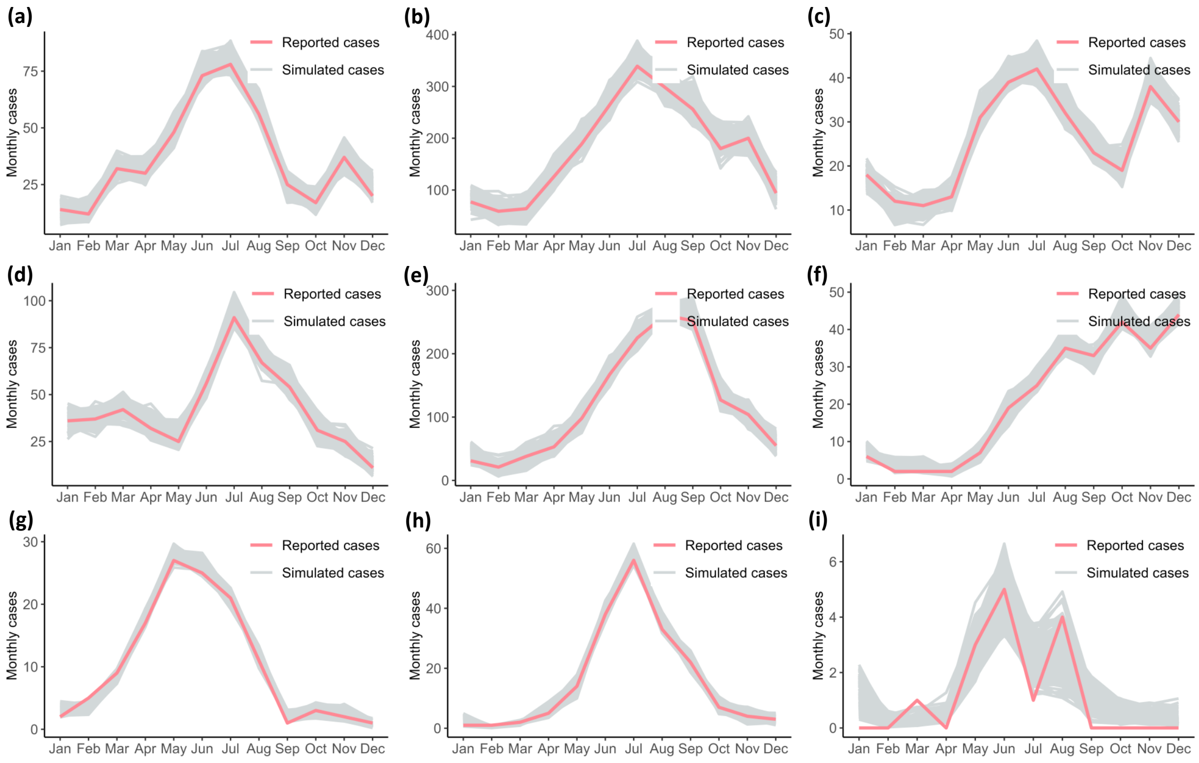


**Figure S1. Model fitting to monthly DHF surveillance data across all province-year combinations.** Results of the model fitting are shown for Rayong (top row), Ratchaburi (middle row), and Phrae (bottom row). Columns from left to right correspond to the years 2006, 2015 and 2017. The pink line shows observed monthly DHF cases. Grey lines represent the ensemble of simulated monthly DHF cases from 200 parameter sets (particles) retained in the final generation of the ABC-SMC algorithm, with each line corresponding to one parameter combination that satisfied the acceptance criterion (RMSE < 14.5% of mean monthly cases). The simulated DHF cases were calculated as the monthly sum of secondary dengue infections multiplied by the estimated reporting proportion. Note that panel (b) (Rayong, 2015) is also presented in the main text as **Figure 3**.

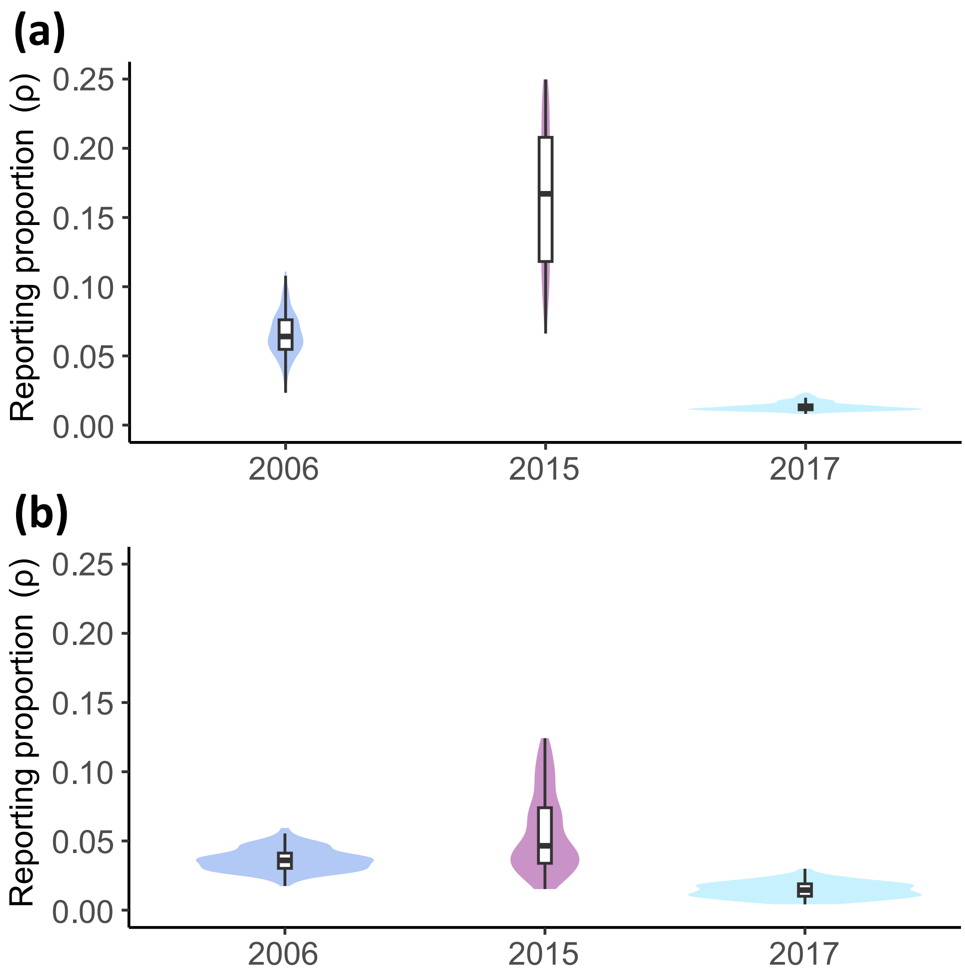


**Figure S2. Posterior distributions of estimated reporting proportions for (a) Ratchaburi and (b) Phrae across three years (2006, 2015, 2017).** Violin plots display the distribution of reporting proportion estimates from 200 parameter sets retained in the final generation of ABC-SMC for each year. The reporting proportion represents the fraction of secondary dengue infections that progress to DHF and are captured by the surveillance system. The width of each violin indicates the kernel density estimate of the posterior distribution, with embedded box plots showing the median (horizontal line), interquartile range (box), and 95% credible intervals (whiskers).

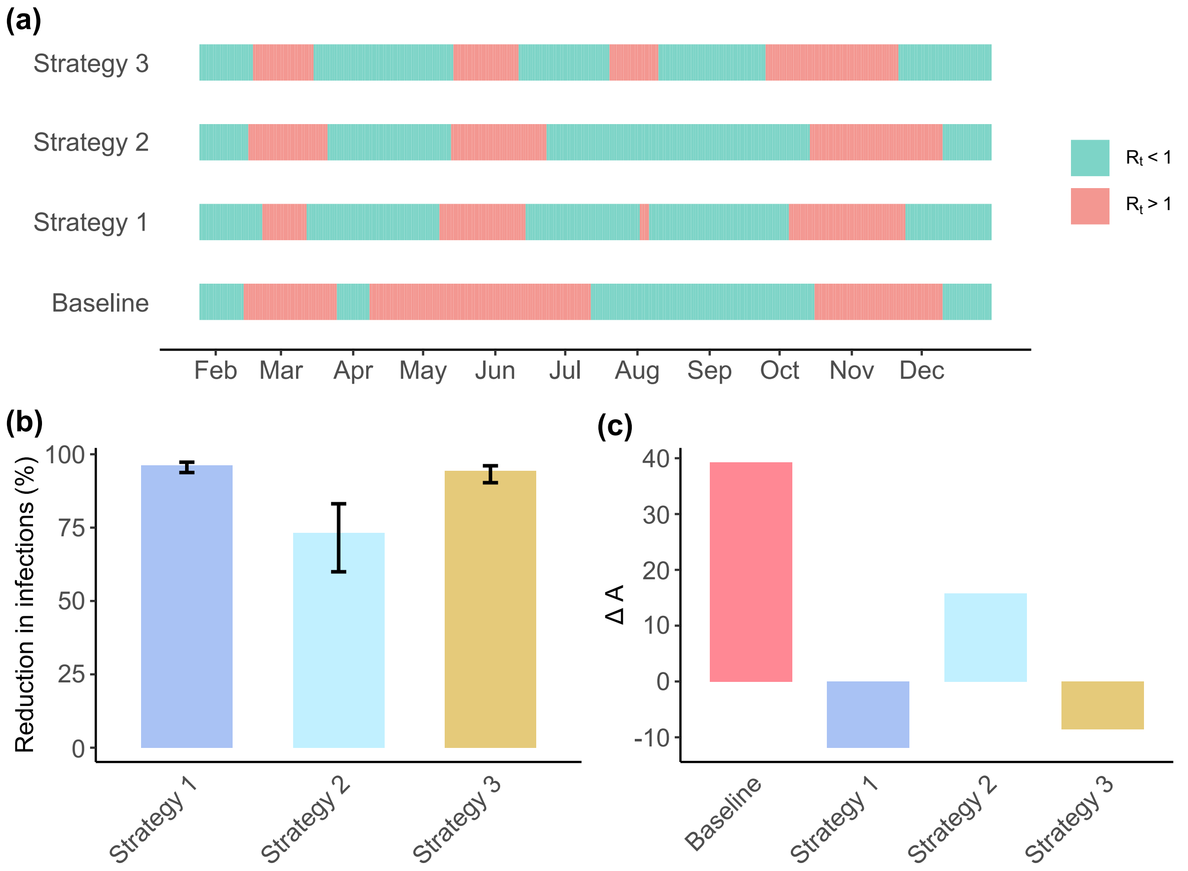


**Figure S3. Quantitative assessment of vector control effectiveness on dengue epidemic dynamics in Rayong province, 2006.** (a) Time-varying effective reproduction number ($R_t$) estimated using EpiEstim with a weekly sliding window and gamma-distributed generation time (mean: 18.2 days, SD: 6.1 days). Red shading indicates periods of epidemic growth ($R_t > 1$) and green shading indicates epidemic decline ($R_t < 1$). Strategy 1 represents bite prevention, while Strategy 2 and Strategy 3 represent the use of larvicides and adulticides, respectively. (b) Percentage reduction in cumulative dengue infections relative to baseline, calculated from 200 parameter sets retained from ABC-SMC. Error bars represent 95% credible intervals derived from the posterior distribution. (c) Net epidemic intensity ($\Delta A$) calculated as the difference between areas above and below the $R_t = 1$ threshold. Negative values indicate successful epidemic suppression, with larger absolute values representing stronger control.

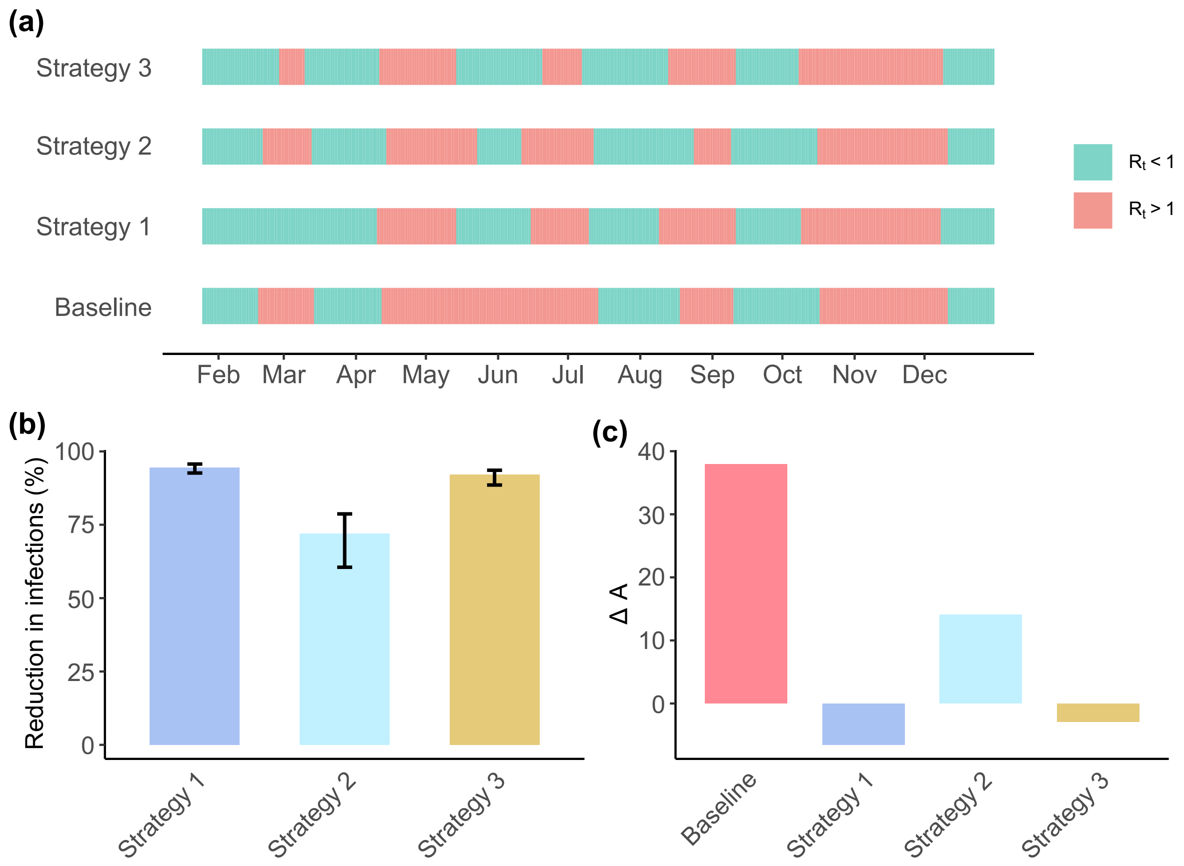


**Figure S4. Quantitative assessment of vector control effectiveness on dengue epidemic dynamics in Rayong province, 2017.** (a) Time-varying effective reproduction number ($R_t$) estimated using EpiEstim with a weekly sliding window and gamma-distributed generation time (mean: 18.2 days, SD: 6.1 days). Red shading indicates periods of epidemic growth ($R_t > 1$) and green shading indicates epidemic decline ($R_t < 1$). Strategy 1 represents bite prevention, while Strategy 2 and Strategy 3 represent the use of larvicides and adulticides, respectively. (b) Percentage reduction in cumulative dengue infections relative to baseline, calculated from 200 parameter sets retained from ABC-SMC. Error bars represent 95% credible intervals derived from the posterior distribution. (c) Net epidemic intensity ($\Delta A$) calculated as the difference between areas above and below the $R_t = 1$ threshold. Negative values indicate successful epidemic suppression, with larger absolute values representing stronger control.

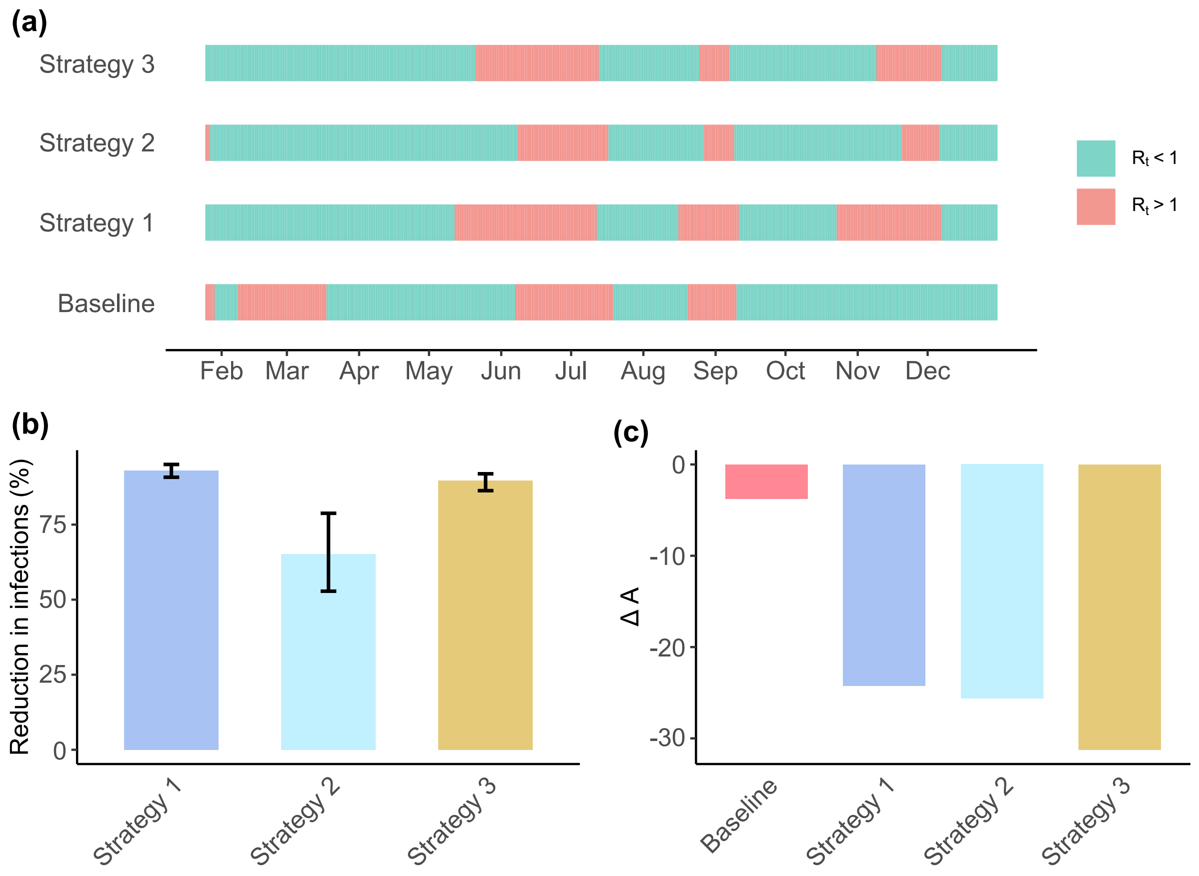


**Figure S5. Quantitative assessment of vector control effectiveness on dengue epidemic dynamics in Ratchaburi province, 2006.** (a) Time-varying effective reproduction number ($R_t$) estimated using EpiEstim with a weekly sliding window and gamma-distributed generation time (mean: 18.2 days, SD: 6.1 days). Red shading indicates periods of epidemic growth ($R_t$ > 1) and green shading indicates epidemic decline ($R_t$ < 1). Strategy 1 represents bite prevention, while Strategy 2 and Strategy 3 represent the use of larvicides and adulticides, respectively. (b) Percentage reduction in cumulative dengue infections relative to baseline, calculated from 200 parameter sets retained from ABC-SMC. Error bars represent 95% credible intervals derived from the posterior distribution. (c) Net epidemic intensity ($\Delta A$) calculated as the difference between areas above and below the $R_t$ = 1 threshold. Negative values indicate successful epidemic suppression, with larger absolute values representing stronger control.

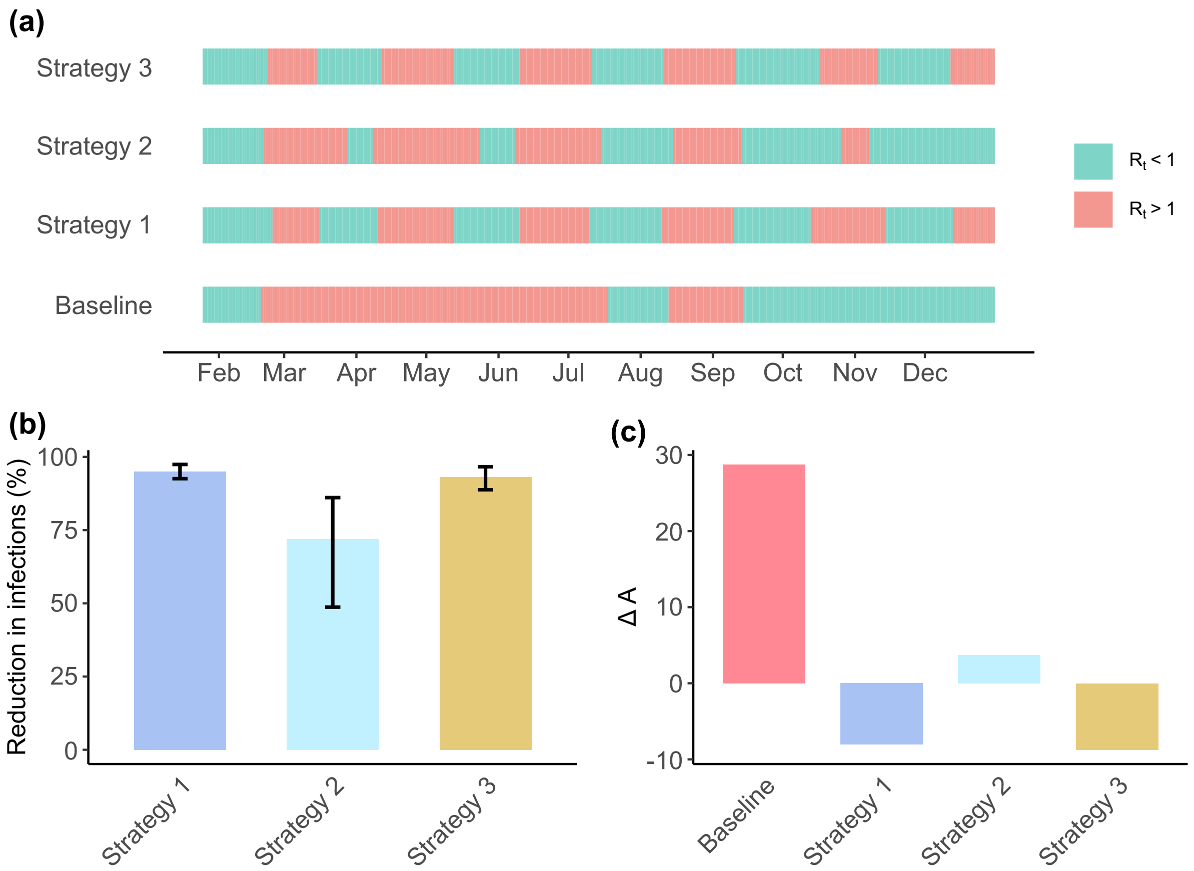


**Figure S6. Quantitative assessment of vector control effectiveness on dengue epidemic dynamics in Ratchaburi province, 2015.** (a) Time-varying effective reproduction number ($R_t$) estimated using EpiEstim with a weekly sliding window and gamma-distributed generation time (mean: 18.2 days, SD: 6.1 days). Red shading indicates periods of epidemic growth ($R_t > 1$) and green shading indicates epidemic decline ($R_t < 1$). Strategy 1 represents bite prevention, while Strategy 2 and Strategy 3 represent the use of larvicides and adulticides, respectively. (b) Percentage reduction in cumulative dengue infections relative to baseline, calculated from 200 parameter sets retained from ABC-SMC. Error bars represent 95% credible intervals derived from the posterior distribution. (c) Net epidemic intensity ($\Delta A$) calculated as the difference between areas above and below the $R_t = 1$ threshold. Negative values indicate successful epidemic suppression, with larger absolute values representing stronger control.

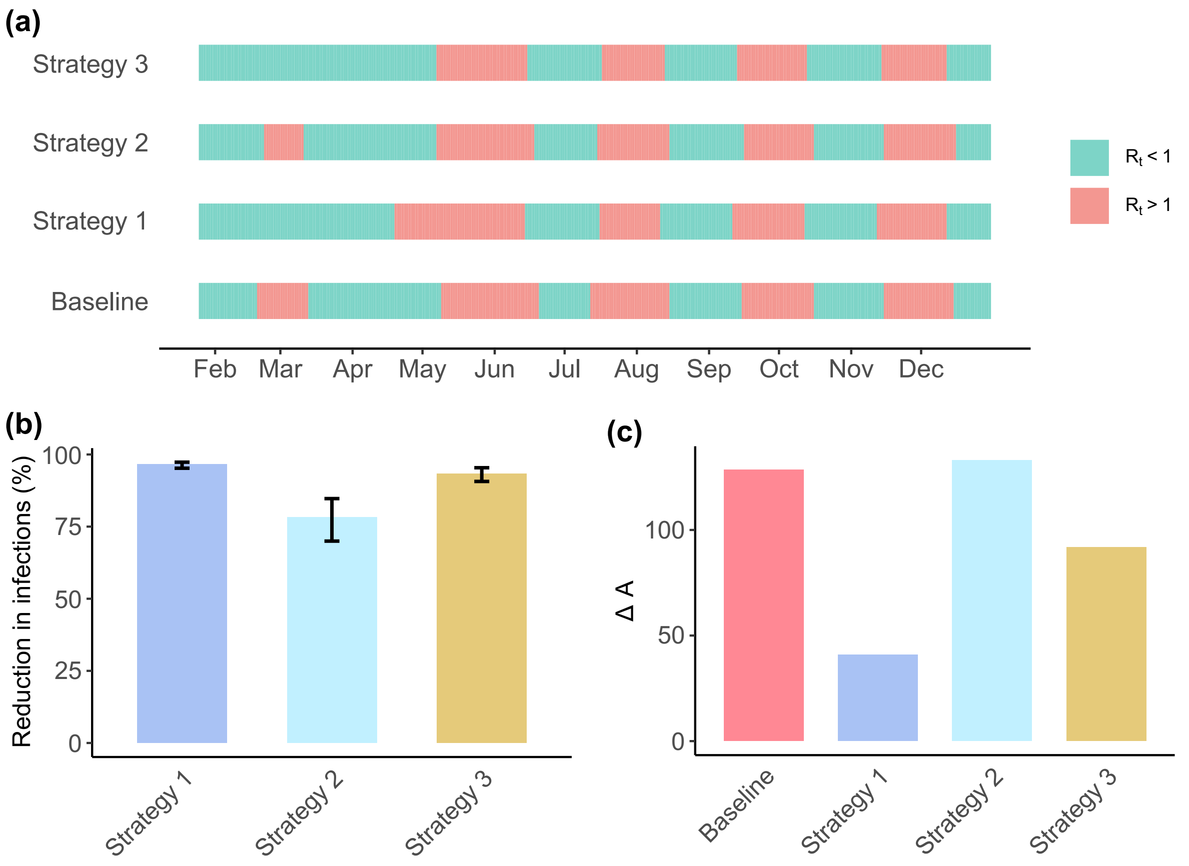


**Figure S7. Quantitative assessment of vector control effectiveness on dengue epidemic dynamics in Ratchaburi province, 2017.** (a) Time-varying effective reproduction number ($R_t$) estimated using EpiEstim with a weekly sliding window and gamma-distributed generation time (mean: 18.2 days, SD: 6.1 days). Red shading indicates periods of epidemic growth ($R_t > 1$) and green shading indicates epidemic decline ($R_t < 1$). Strategy 1 represents bite prevention, while Strategy 2 and Strategy 3 represent the use of larvicides and adulticides, respectively. (b) Percentage reduction in cumulative dengue infections relative to baseline, calculated from 200 parameter sets retained from ABC-SMC. Error bars represent 95% credible intervals derived from the posterior distribution. (c) Net epidemic intensity ($\Delta A$) calculated as the difference between areas above and below the $R_t = 1$ threshold. Negative values indicate successful epidemic suppression, with larger absolute values representing stronger control.

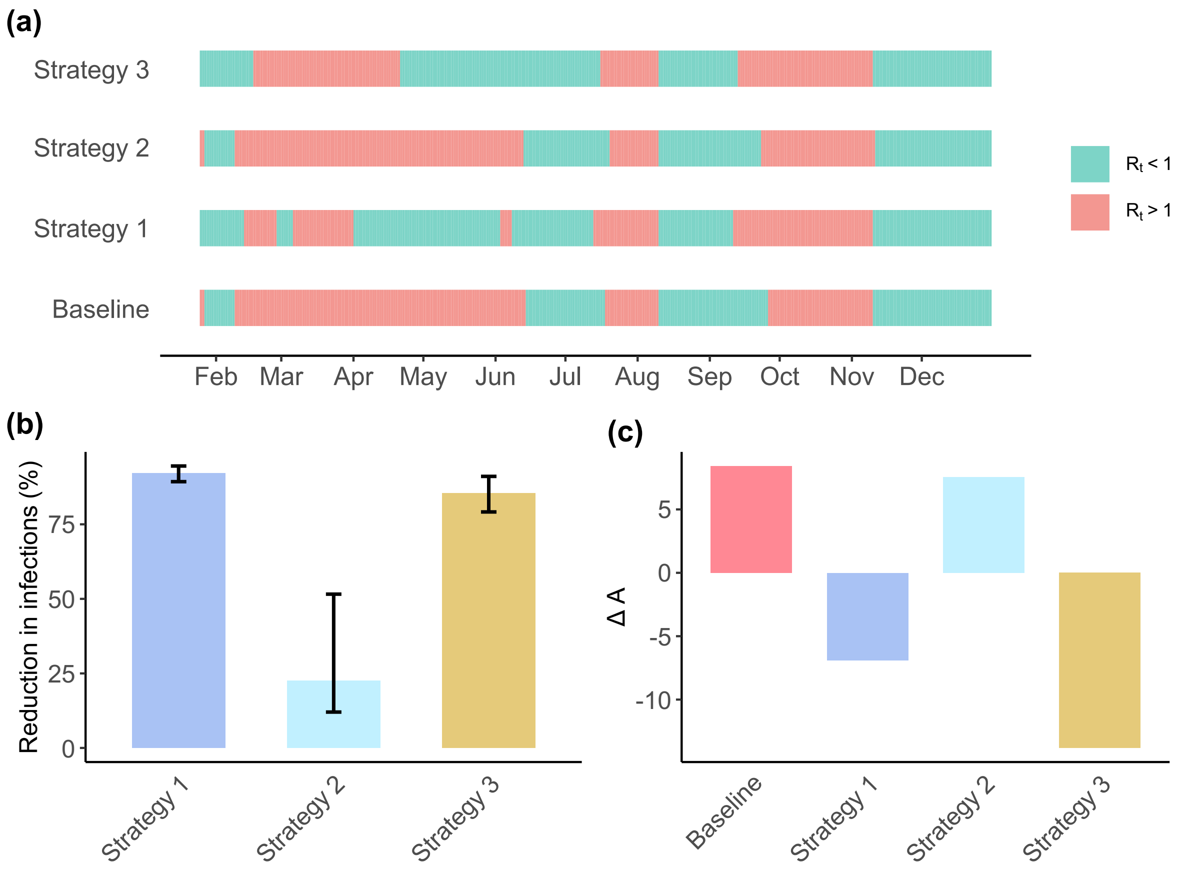


**Figure S8. Quantitative assessment of vector control effectiveness on dengue epidemic dynamics in Phrae province, 2006.** (a) Time-varying effective reproduction number ($R_t$) estimated using EpiEstim with a weekly sliding window and gamma-distributed generation time (mean: 18.2 days, SD: 6.1 days). Red shading indicates periods of epidemic growth ($R_t > 1$) and green shading indicates epidemic decline ($R_t < 1$). Strategy 1 represents bite prevention, while Strategy 2 and Strategy 3 represent the use of larvicides and adulticides, respectively. (b) Percentage reduction in cumulative dengue infections relative to baseline, calculated from 200 parameter sets retained from ABC-SMC. Error bars represent 95% credible intervals derived from the posterior distribution. (c) Net epidemic intensity ($\Delta A$) calculated as the difference between areas above and below the $R_t$ = 1 threshold. Negative values indicate successful epidemic suppression, with larger absolute values representing stronger control.

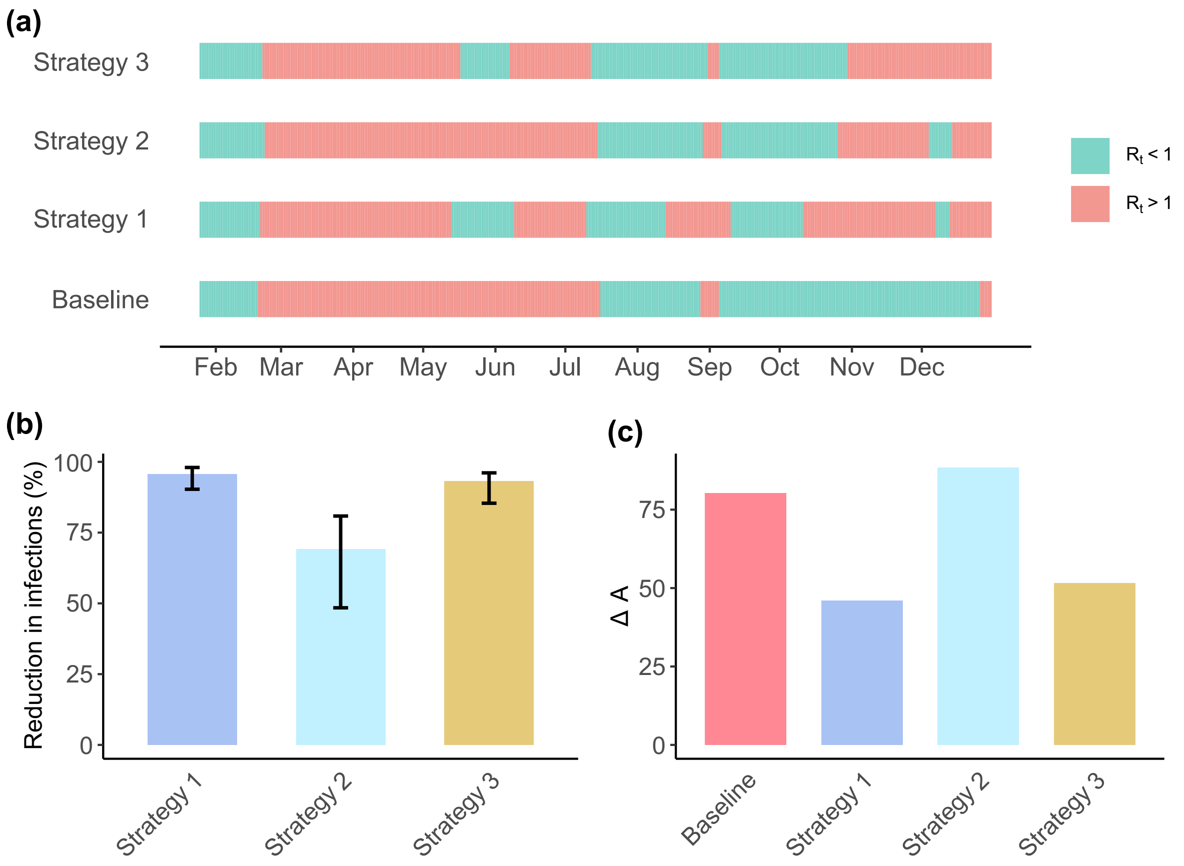


**Figure S9. Quantitative assessment of vector control effectiveness on dengue epidemic dynamics in Phrae province, 2015.** (a) Time-varying effective reproduction number ($R_t$) estimated using EpiEstim with a weekly sliding window and gamma-distributed generation time (mean: 18.2 days, SD: 6.1 days). Red shading indicates periods of epidemic growth ($R_t > 1$) and green shading indicates epidemic decline ($R_t < 1$). Strategy 1 represents bite prevention, while Strategy 2 and Strategy 3 represent the use of larvicides and adulticides, respectively. (b) Percentage reduction in cumulative dengue infections relative to baseline, calculated from 200 parameter sets retained from ABC-SMC. Error bars represent 95% credible intervals derived from the posterior distribution. (c) Net epidemic intensity ($\Delta A$) calculated as the difference between areas above and below the $R_t = 1$ threshold. Negative values indicate successful epidemic suppression, with larger absolute values representing stronger control.

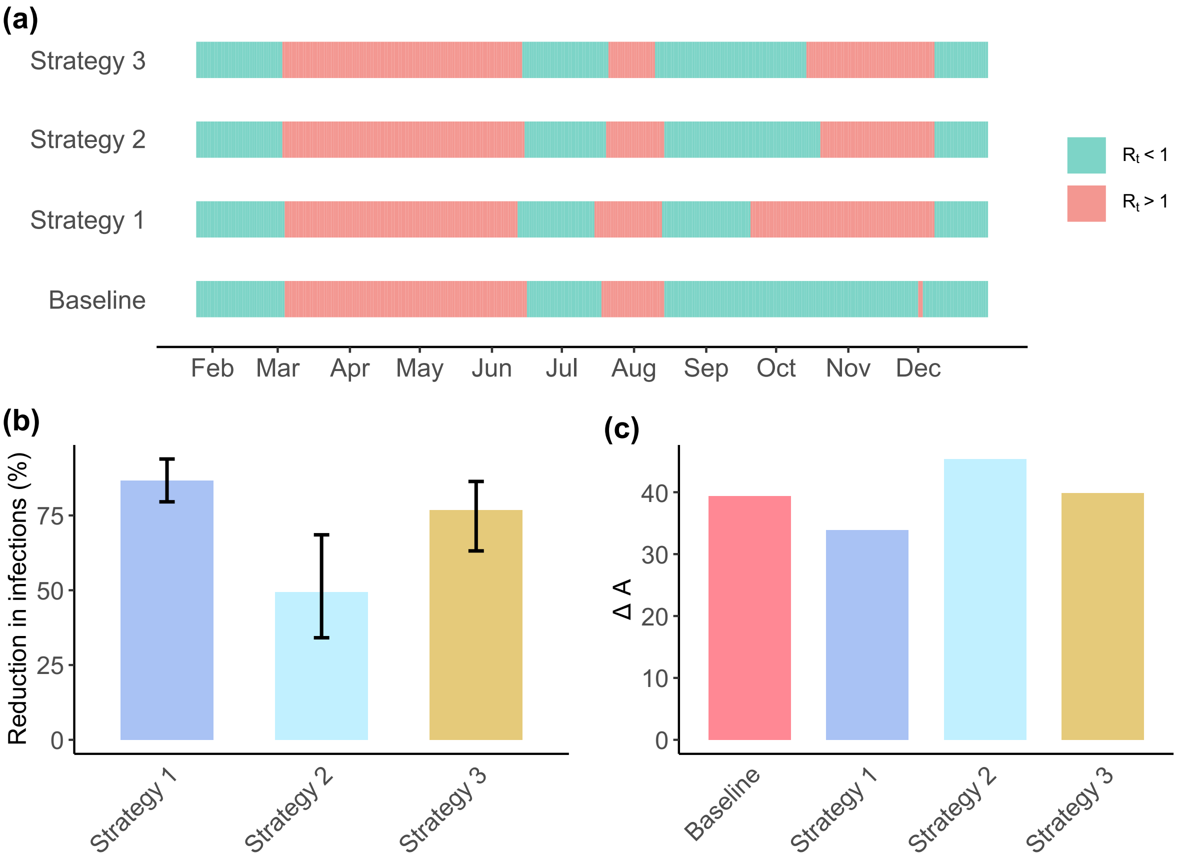


**Figure S10. Quantitative assessment of vector control effectiveness on dengue epidemic dynamics in Phrae province, 2017.** (a) Time-varying effective reproduction number ($R_t$) estimated using EpiEstim with a weekly sliding window and gamma-distributed generation time (mean: 18.2 days, SD: 6.1 days). Red shading indicates periods of epidemic growth ($R_t$ > 1) and green shading indicates epidemic decline ($R_t$ < 1). Strategy 1 represents bite prevention, while Strategy 2 and Strategy 3 represent the use of larvicides and adulticides, respectively. (b) Percentage reduction in cumulative dengue infections relative to baseline, calculated from 200 parameter sets retained from ABC-SMC. Error bars represent 95% credible intervals derived from the posterior distribution. (c) Net epidemic intensity ($\Delta A$) calculated as the difference between areas above and below the $R_t$ = 1 threshold. Negative values indicate successful epidemic suppression, with larger absolute values representing stronger control.
